# The Heterogeneous Effect of High PEEP strategies on Survival in Acute Respiratory Distress Syndrome: preliminary results of a data-driven analysis of randomized trials

**DOI:** 10.1101/2025.01.23.25320649

**Authors:** J.M. Smit, J.H. Krijthe, J. Van Bommel, D.S. Sulemanji, J. Villar, F. Suárez-Sipmann, R.L. Fernández, F.G. Zampieri, I.S. Maia, A.B. Cavalcanti, M. Briel, M.O. Meade, Q. Zhou, R.G. Brower, P. Sinha, B. Bartek, C.S. Calfee, A. Mercat, J-C Richard, L. Brochard, A. Serpa Neto, C. Hodgson, E.N. Baedorf-Kassis, D. Talmor, D. Gommers, M.E. Van Genderen, M.J.T. Reinders, A.H. Jonkman

**Affiliations:** Department of Intensive Care, Erasmus MC - University Medical Center Rotterdam, Rotterdam, The Netherlands; Pattern Recognition & Bioinformatics group, Delft University of Technology, Delft, The Netherlands; Critical Care Section, Department of Medicine, Aga Khan University Hospital, Nairobi, Kenya; CIBER de Enfermedades Respiratorias, Instituto de Salud Carlos III, Madrid, Spain; Research Unit at Hospital Universitario Dr, Negrín, Fundación Canaria Instituto de Investigación Sanitaria de Canarias, Las Palmas de Gran Canaria, Spain; Li Ka Shing Knowledge Institute, St. Michael’s Hospital, Toronto, Canada; Faculty of Health Sciences, Universidad del Atántico Medio, Las Palmas, Spain; Intensive Care Unit, Hospital Universitario de la Princesa, Madrid, Spain; Department of Critical Care Medicines, University of Alberta, Edmonton, Canada; Hcor Research Institute, São Paulo, Brazil; CLEAR Methods Center, Division of Clinical Epidemiology, Department of Clinical Research, University of Basel and University Hospital Basel, Basel, Switzerland; Department of Health Research Methodology, Evidence, and Impact, McMaster University, Hamilton, Canada; Division of Critical Care, Department of Medicine, McMaster University; Pulmonary and Critical Care Medicine, Johns Hopkins University School of Medicine; Department of Anesthesiology, Washington University School of Medicine, St. Louis, MO, USA; Department of Medicine, Division of Pulmonary, Critical Care, Allergy and Sleep Medicine; University of California, San Francisco; San Francisco, CA; Department of Anesthesia; University of California, San Francisco; San Francisco, CA; Médecine Intensive - Réanimation, VentLab, CHU D’Angers. Université d’Angers. Angers, France; Keenan Centre for Biomedical Research, Li Ka Shing Knowledge Institute, Unity Health Toronto, Toronto, Ontario, Canada; Interdepartmental Division of Critical Care Medicine, University of Toronto, Toronto, Ontario, Canada; Australian and New Zealand Intensive Care Research Centre (ANZIC-RC), School of Public Health and Preventive Medicine, Monash University, Melbourne, Australia; Department of Intensive Care, Austin Hospital, Melbourne, Australia; Department of Critical Care, Melbourne Medical School, University of Melbourne, Austin Hospital, Melbourne, Australia; Department of Critical Care Medicine, Hospital Israelita Albert Einstein, Sao Paulo, Brazil; Intensive Care Unit, Alfred Health, Melbourne, Australia; Department of Anaesthesia, Critical Care, and Pain Medicine, Beth Israel Deaconess Medical Center and Harvard Medical School, Boston, MA USA

**Author notes:** **Corresponding author:** Jim Smit, Erasmus MC – University Medical Center Rotterdam, Department of Intensive Care (internal postal address: Room Ne-411) Doctor Molewaterplein 40, 3015 GD Rotterdam, the Netherlands.

## Abstract

**Background:** Mixed trial results suggest that some ventilated patients with acute respiratory distress syndrome (ARDS) benefit from high PEEP while others may be harmed, indicating heterogeneity of treatment effect (HTE). This study applies data-driven predictive approaches to uncover HTE and re-examines previously hypothesized HTE. This manuscript serves as a pre-registration of planned external validation of our trained models.

**Methods:** We identified eight randomized trials, and obtained individual patient data (IPD) from three of them (ALVEOLI, LOVS, EXPRESS), as our train cohort. We used effect modelling to predict individualized treatment effects (predicted 28-day mortality risk difference between PEEP strategies) across patient subgroups stratified by observed tertiles (≤8 cmH_2_O, 9–11 cmH_2_O, ≥12 cmH_2_O). Candidate effect modelling methods included meta-learners and technique-specific methods. Optimal methods were selected through ‘leave-one-trial-out’ cross-validation, evaluating the methods’ performances in each PEEP tertile using AUC-benefit. We trained final models using the best performing methods implemented with or without forward selection (which yielded sufficient AUC-benefit), and additional final models by selecting the variables that yielded consistency in the forward selections performed in the cross validation, if any. We further evaluated earlier hypothesized HTE comparing (1) patients with baseline PaO2/FiO2 ≤ 200 versus > 200 mmHg, and (2) patients with hypoinflammatory versus hyperinflammatory subphenotypes.

**Preliminary findings:** In the lower PEEP tertile (≤8 cmH_2_O), an X-learner implemented without, and an S-learner implemented with forward selection (both with flexible base learners), yielded the highest AUC benefits and were used to train final models. In the high PEEP tertile (≥12 cmH_2_O), only the causal forest implemented with forward selection yielded an AUC benefit exceeding zero. Respiratory-system compliance (C_RS_) was consistently selected in the forward selections of cross validation, and was used to train an extra final causal forest model, with predicted effects shifting from harm to benefit for C_RS_ 26.5 mL/cmH_2_O or higher.

Higher PEEP benefited patients with baseline PaO_2_/FiO_2_ ≤200 mmHg (OR 0.80, 95% CI 0.66–0.98), incurred harm among those with PaO_2_/FiO_2_ >200 mmHg (OR 1.74, 95% CI 1.02–2.98; interaction P=0.01). This HTE was strongest when PaO_2_/FiO_2_ was measured at low PEEP (≤8 cmH_2_O), reduced at mid-level PEEP (9–11 cmH_2_O), and negligible at high PEEP (≥12 cmH_2_O). A second-order interaction showed significant heterogeneity of HTE (ie, second-order heterogeneity) across PEEP tertiles (P=0.03).

**Preliminary Conclusions:** Our preliminary findings indicated that baseline C_RS_ ≥ 26.5 mL/cmH_2_O predicts benefit, while C_RS_ < 26.5 mL/cmH_2_O predicts harm from high PEEP when C_RS_ is measured at high baseline PEEP (≥12 cmH_2_O). Similarly, baseline PaO_2_/FiO_2_ ≤ 200 mmHg predicts benefit, while PaO_2_/FiO_2_ > 200 mmHg predicts harm from high PEEP when PaO_2_/FiO_2_ is measured at a low baseline PEEP (≤8 cmH_2_O). Using data from the LOVS trial, we investigated HTE for high PEEP between hypo- and hyperinflammatory subphenotypes but found none, despite significant HTE observed earlier in the ALVEOLI trial.

## 1. Introduction

First conceptualized by Lachmann^1^ and introduced in randomized controlled trials (RCTs) by Amato et al.,^2,3^ the ‘open lung approach’ (OLA) aims to maximize lung aeration in mechanically ventilated patients with acute respiratory distress syndrome (ARDS) by performing recruitment manoeuvres (RMs) to reverse atelectasis and then applying high levels of positive end-expiratory pressure (PEEP) to keep alveoli open. The first three RCTs on this topic, which compared lower versus higher PEEP strategies (with or without RMs),^4–6^ were inconclusive on crude mortality. In 2010, an individualized patient data meta-analysis of these trials by Briel et al.^7^ revealed significant heterogeneity of treatment effect (HTE), suggesting that only patients with baseline PaO_2_/FiO_2_ ≤ 200 mmHg (currently known as ‘moderate-severe ARDS’) benefitted from higher PEEP (ie, “Briel’s hypothesis”). This led to subsequent (and upcoming) RCTs^8–13^ *excluding* patients with baseline PaO_2_/FiO_2_ > 200 mmHg for examining the effects of higher PEEP approached. However, despite expectations of positive results, the largest RCT^9^ comparing lower vs higher PEEP reported a significant mortality *increase* in the higher PEEP group. Following this unexpected and disappointing result, which may have different explanations,^14^ some investigators have proposed that the research community should consider discontinuing efforts on OLA and shift focus to other strategies.^15^ It remains unexamined, however, what the effect is of the PEEP setting at which the baseline PaO_2_/FiO_2_ was measured on the previously reported effect heterogeneity. This is important to consider, as it is well-known that the set PEEP level affects various respiratory variables such as PaO_2_/FiO_2_ and respiratory-system compliance (C_RS_).^16^

Our study has two aims: (1) to conduct an exploratory predictive HTE analysis^17^ to identify patients likely to benefit from higher PEEP based on their baseline characteristics, and (2) to reassess prior hypotheses on HTE for higher versus lower PEEP, specifically comparing patients with baseline PaO_2_/FiO_2_ ≤ 200 versus > 200 mmHg,^7^ and those with hypoinflammatory versus hyperinflammatory subphenotypes.^18^ A key aspect of our approach is stratifying patients based on the PEEP setting at the time where baseline variables were measured.

## 2. Methods

### 2.1 Trial Selection, Data Collection and Study endpoint

In accordance with a recent aggregate data meta-analysis by Neto et al.,^19^ we included RCTs that compared strategies to determine PEEP levels in adult patients with ARDS, which demonstrated a difference in achieved PEEP levels between the groups, and were published after 2000, when the low tidal volume ventilation trial was published.^20^ Serpa-Neto et al.,^19^ who searched MEDLINE, EMBASE and Cochrane Central Register of Controlled Trials from 1996 to March 1^st^ 2020, identified eight eligible trials. For three of these RCTs (ie, the ALVEOLI,^4^ LOVS^5^ and EXPRESS^6^ trials), we obtained individual patient data (IPD), forming the *train cohort*, whereas the remaining five trials^8–11,21^ will form the *test cohort*. Crucially, we requested the authors of the trials forming the test cohort to provide IPD *after* publishing this preliminary report, ensuring that the IPD of the test cohort could not influence choices in model training or evaluation. Collected IPD included age, sex, clinical parameters, baseline ventilatory and laboratory values. All included RCTs had ethical approval from local institutional Medical Ethics Committees. The primary study endpoint is 28-day all-cause mortality. All patients were analysed in the study group to which they were randomised (intention-to-treat principle).

### 2.2 Explorative predictive HTE analysis

We conducted an explorative predictive HTE analysis, using *effect modelling*, which, as opposed to ‘risk modelling’, aims to predict ‘individualized treatment effects’, conditional on patient baseline characteristics.^17^ We defined the individualized treatment effect as the predicted probability of 28-day mortality given a lower PEEP regime, *minus* the predicted probability of 28-day mortality given a higher PEEP regime (**Figure 1**).

**Figure 1:**
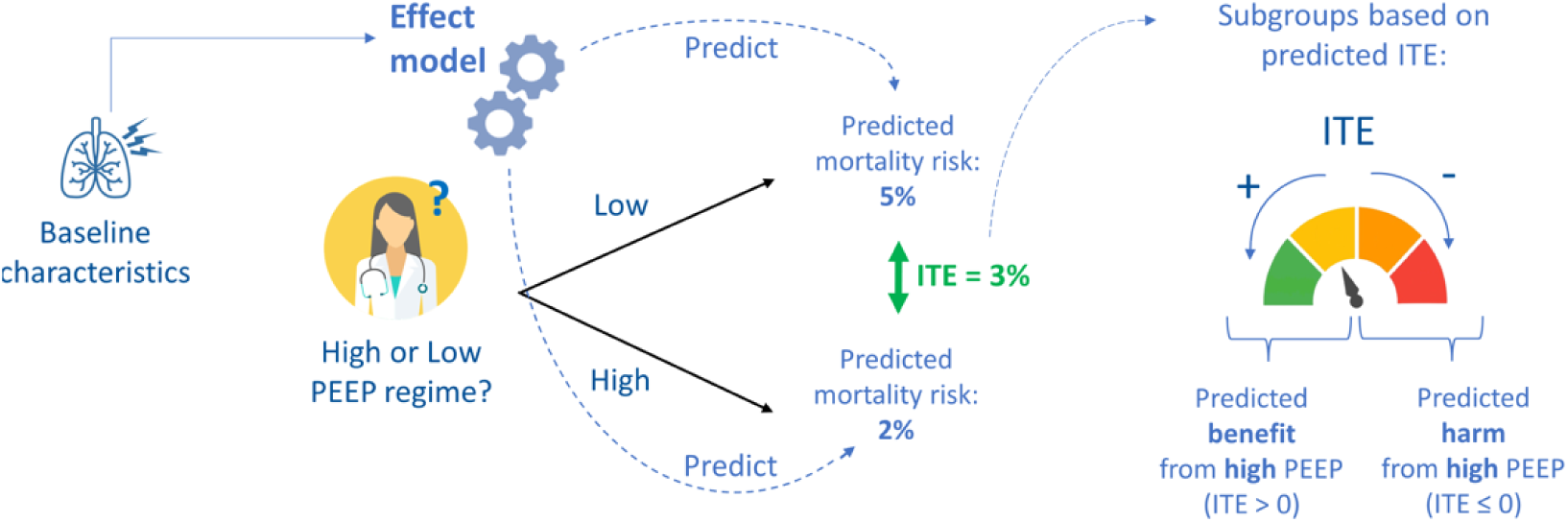
Schematic overview of the prediction of individualized treatment effects through effect modelling. An effect model predicts for every patient, based on baseline characteristic(s), an individualized treatment effect (ITE), ie, predicted probability of 28-day mortality given a lower PEEP regime, *minus* the predicted probability of 28-day mortality given a higher PEEP regime. Using these predicted ITEs, patients are classified in two subgroups: those with predicted benefit from high PEEP, and those with predicted harm from high PEEP.

The explorative analysis consists of four steps (**Figure 2**):

**Figure 2:**
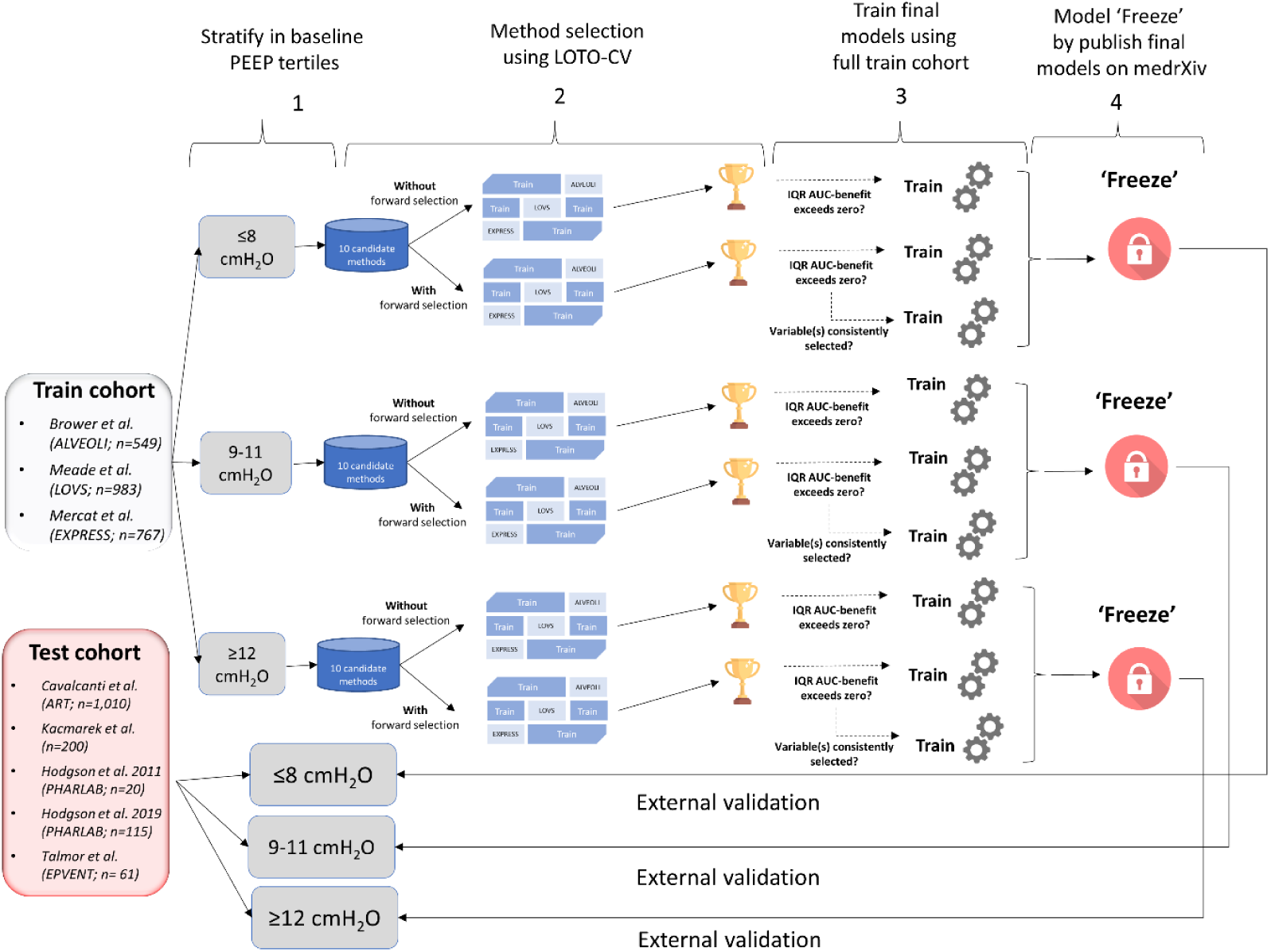
Schematic overview of the explorative predictive HTE analysis through effect modelling: 1) the stratification of the train cohort into baeline PEEP tertiles; 2) among 10 candidate methods, each implemented with, and without forward selection, select the method leading to the highest discriminative performance (ie, AUC-benefit) resulting from leave-one-trial-out cross-validation (LOTO-CV) procedures in each PEEP tertile; 3) training of final models using best performing methods (if any) in each PEEP tertile, using the full train cohort (highlighted in green); 4) ‘freeze’ models by pre-registering these on medRxiv and externally validating these in the patients from the corresponding PEEP tertiles in the test cohort.

#### Step 1: Stratifying patients by set baseline PEEP

Because the set PEEP affects various blood-gas based and respiratory variables (such as PaO_2_/FiO_2_ and C_RS_),^16^ we adopted the principle to only train and evaluate effect models within patient subgroups, such that baseline characteristics were measured at a similar PEEP level. The baseline PEEP distributions among patients in the train cohort showed three peaks, centred around 5, 10 and 15 cmH_2_O (**Figure 3**). Hence, we first divided all patients in three groups, based on baseline PEEP tertiles as observed in the train cohort: ≤8 cmH_2_O, 9-11 cmH_2_O and ≥12 cmH_2_O. We trained and evaluated effect models separately within these subgroups.

**Figure 3:**
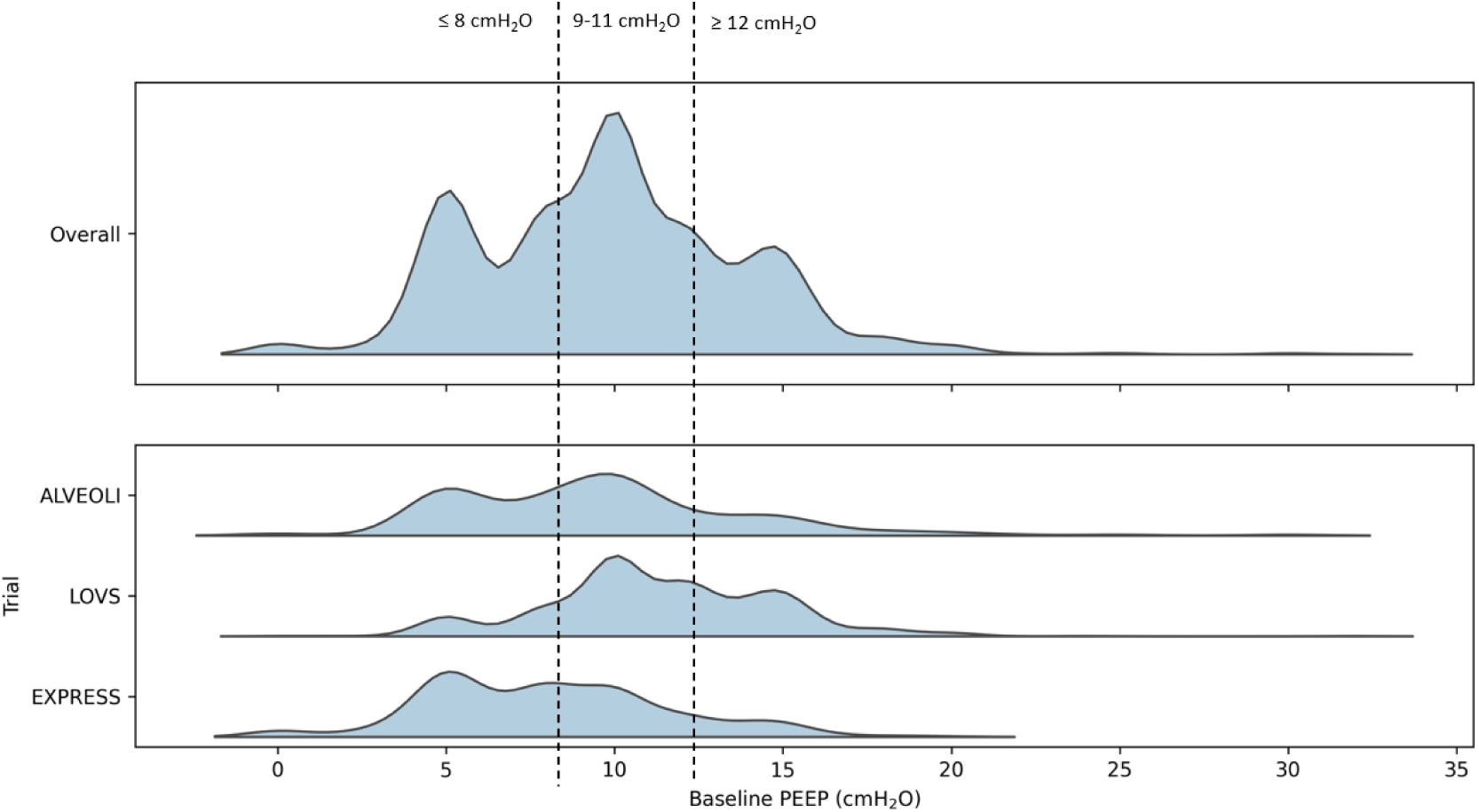
Distribution of the baseline positive end-expiratory pressure (PEEP) in the whole train cohort, and in each trial of the train cohort separately. During the explorative, predictive HTE analysis, the patients are split based on the observed tertiles in the full train cohort.

#### Step 2: Method selection

We compared several effect modelling methods (ie, ‘candidate methods’) with varying degrees of flexibility. Specifically, we experimented with various ‘meta-learners’, which are model-agnostic algorithms that break down the estimation of individualized treatment effects into multiple sub-problems, each solvable with any supervised machine learning (ML) model, known as the ‘base model’.^22^ We implemented ‘S-learners’, consisting of a single model that includes treatment as a variable; ‘T-learners’, consisting of two separate models, one trained on the treatment group and one on the control group; and the more complex ‘X-learners’^23^ and ‘R-learners’.^24^ We implemented each meta-learner using either a linear (Lasso logistic regression) or a more flexible (Light Gradient-Boosting Machine [LightGBM]) base model, referred to as the ‘S/T/X/R-Lasso’ and ‘S/T/X/R-GBM’ models, respectively. Additionally, we examined two candidate methods which are (in contrast to meta-learners) specific to certain ML techniques: the Tian method,^25^ based on Lasso logistic regression, and the causal forest, which is a generalization of the random forest, designed specifically for the prediction of individualized treatment effects,^26^ totalling 10 different candidate methods. As we only received data from the three trials^4–6^ (the train cohort; see step 2.1), we could only use these data to select the optimal effect modelling method in each PEEP tertile. This method selection process consisted of several sub-steps.

#### Step 2.1 A-priori variable selection

A priori, we selected baseline variables (including demographics, vital signs, blood gas based variables and respiratory variables) which were available in each trial of the train cohort, as well as in the ART trial,^9^ as this trial comprises 72% of the test cohort data. In the ART trial,^9^ patients first received mechanical ventilation based on the lower PEEP-FiO_2_ ARDSnet table,^4^ during which baseline respiratory variables were collected. Subsequently, FiO_2_ was set to 100% and PEEP to 10 cmH_2_O for all patients, and baseline blood gas measurements were collected. Hence, respiratory variables (such as C_RS_) were measured at *varying* PEEP levels among patients, whereas blood gas-based variables (such as PaO_2_/FiO_2_) were measured only in the middle PEEP tertile. To stick with our principle from step 1 for patients in the lower and higher PEEP tertiles, we only trained models after a-priori excluding blood gas-based variables. For complete lists of the a-priori selections for each PEEP tertile, see **Supplementary Table S1**.

#### Step 2.2 Data pre-processing

Before training the effect models, we first normalized the data by centring and scaling each variable, and addressed missing values using the K-Nearest-Neighbour (KNN; with K neighbours = 5) imputation algorithm. After imputing the missing values, the variables were rescaled to the original scales, and normalized again by centring and scaling each variable, ensuring input data with zero-mean and unit variance before training any effect model.

#### Step 2.3 Leave-one-trial-out cross-validation (LOTO-CV)

In each PEEP tertile, we performed a ‘leave-one-trial-out’ cross-validation (LOTO-CV) procedure within the train cohort, referred to as the ‘outer’ LOTO-CV, to evaluate the discriminative performance in terms of ‘AUC-benefit’ (appendix part 2) of each candidate method. This outer LOTO-CV was as follows (**Supplementary Figure S1**). In each of the three CV ‘folds’, we split the train cohort (three trials) into an inner-train cohort (two trials) and an inner-test cohort (one trial), in which each of the three trials forms the inner-test cohort once. In each CV fold, first the hyperparameters were optimized using a ‘nested’ LOTO-CV (see next substep), an effect model was trained using the inner-train cohort, and individualized treatment effects were predicted for the patients in the inner-test cohort.

After the three CV folds, all predictions were combined, and 200 bootstrap samples were taken from this combined set of predictions. Within each bootstrap sample, the AUC-benefit was calculated for each set of sampled predictions originating from the same CV fold, and we combined these three AUC-benefits by calculating the weighted average (see appendix part 2). Hence, each bootstrap sample resulted in one weighted-average AUC-benefit. From the 200 weighted-average AUC-benefits, we calculated the median. In each PEEP tertile, we selected the candidate method leading to the highest median AUC-benefit. However, the selected method was only considered if the inter-quartile-range (IQR) of the bootstrapped weighted-average AUC-benefits exceeded zero. Otherwise, no method was selected.

#### Step 2.4 Hyperparameter optimization through ‘nested’ LOTO-CV

Each candidate method we examined has specific hyperparameters. For each candidate method, we optimized a method-specific set of hyperparameters using an exhaustive grid search (see **Supplementary Table S2** for the searched grids). In each CV fold of the outer LOTO-CV, we performed ‘nested’ LOTO-CVs, to evaluate the discriminative performance of each unique combination of hyperparameters. This nested LOTO-CV is as follows (**Supplementary Figure S1**). In each of the two nested CV ‘folds’, we split the inner train cohort (two trials) into an inner-inner train cohort (one trial) and an inner-inner test cohort (one trial), in which each of the two trials forms the inner-inner test cohort once. In each nested CV fold, an effect model was trained using the inner-inner train cohort, implemented with the combination of hyperparameters as considered in the grid search, and individualized treatment effects were predicted for the patients in the inner-inner test cohort. After the two nested CV folds, all predictions were combined and the median from the bootstrapped weighted-average AUC-benefits is calculated in the same fashion as for the outer LOTO-CV (see previous substep). We selected the combination of hyperparameters leading to the highest median AUC-benefit.

#### Step 2.5 Forward selection

We hypothesized that further variable selection (ie, after a-priori selection) could reduce overfitting and improve model generalizability to new trial datasets. Therefore, in each PEEP tertile, we repeated the outer LOTO-CV procedure (step 2.3) of each candidate method, but now implementing these with a forward selection, prior to the hyperparameter optimization. Again, we selected for each PEEP tertile the candidate method leading to the highest median AUC-benefit resulting from the outer LOTO-CV procedure, only if the IQR of the bootstrapped AUC-benefits exceeded zero (**Supplementary Figure S1**).

This forward selection procedure was as follows (**Supplementary Figure S2**). In each CV fold of the outer LOTO-CV, prior to the hyperparameter optimization, all a-priori selected variables now form the ‘candidate variables’. For each candidate variable, we performed ‘nested’ LOTO-CVs to evaluate the discriminative performance. Again, in each of the two nested CV ‘folds’, we split the inner train cohort (two trials) into an inner-inner train cohort (one trial) and an inner-inner test cohort (one trial), and trained an effect model using the inner-inner train cohort, implemented with only the candidate variable (and using default hyperparameters), and Individualized treatment effects were predicted for the patients in the inner-inner test cohort. After the two nested CV folds, all predictions were combined and the median from the bootstrapped weighted-average AUC-benefits is calculated in the same fashion as for the outer LOTO-CV (see step 2.4), and we selected the variable leading to the highest median AUC-benefit, removing this one from the candidate variables. Subsequently, a new round is started, training effect models using the previously selected variable, and one of the remaining candidate variables, selecting the variable leading to the largest increase in median AUC-benefit compared to the highest AUC-benefit retrieved in the previous round (if any). This process was repeated until the median AUC did not further increase, or all candidate variables were selected. After this, hyperparameter optimization was performed again, as described in step 2.4.

#### Step 2.6 Training constraint

We hypothesized *qualitative* HTE,^27^ meaning that, some patients *benefit* from, and some are *harmed* by higher PEEP regimes. Hence, for each of the trials included in our training cohort, we assume that there is at least one patient included who benefits from, and at least one patient included who is harmed by higher PEEP strategies. Therefore, we only considered effect models that predict individualized treatment effects indicating benefit (ie, predicted individualized treatment effects > 0) as well as harm (ie, predicted individualized treatment effects ≤ 0; **Figure 1**), each in at least one patient. Consequently, combinations of hyperparameters during the hyperparameter optimization, or candidate variables during the forward selection, which resulted in an effect model that predict either benefit or harm for all patients (in at least one nested CV fold), were not considered. Due to this constraint, it could happen that none of the combinations in the hyperparameter optimization, or none of the candidate variables in the forward selection, were selected. If that occurred in one or more of the outer CV folds, the respective method was not considered at all.

#### Step 3.1: Training of final models

After the optimal effect modelling methods were selected (potentially one without, and one with forward selection, for each PEEP tertile), we trained the ‘final models’ using these selected methods in the data of the full train cohort. The optimization of hyperparameters and forward selection for these final models were performed using the same nested LOTO-CV procedures as used for the method selection (section 2.3), but with the difference that these LOTO-CVs were performed in the full train cohort with three trials (and hence, three LOTO-CV folds) instead of two (**Supplementary Figure S3**).

#### Step 3.2: Training of additional final models

Additionally, we hypothesized that variables which were consistently selected in the forward selection procedures in each fold of the outer LOTO-CV during the method selection, most likely represent effect modifiers which generalize to unseen trial datasets. Hence, in each PEEP tertile, if a final model with forward selection was trained, and if at least one variable was consistently selected in all three CV folds of the outer LOTO-CV during the method selection, we trained another final model, using the same selected method, but rather than performing forward selection, only using the variables that were consistently selected in the LOTO-CV procedure. Hence, maximally nine final models can be trained, ie, for each PEEP tertile, one using the selected method using all a priori selected variables, one using the selected method implemented with forward selection, and one using the same selected method with forward selection, but instead of a forward selection, selecting the variable(s) which were consistently selected in the forward selections across the outer CV folds (**Figure 2**).

#### Step 4: Pre-registering of final models

Each trained final model is pre-registered and will eventually be externally validated in patients in the corresponding PEEP tertiles in the yet to be shared-test cohort.

### 2.3 Evaluation of earlier hypothesized HTE

#### A closer look at Briel’s hypothesis

We first reproduced the findings by Briel and colleagues,^7^ in three steps: (1) stratifying patients in two subgroups: those with baseline PaO_2_/FiO_2_ > 200 mmHg and those with baseline PaO_2_/FiO_2_ ≤ 200 mmHg, (2) estimating relative treatment effects in terms of odds ratios (ORs) for these subgroups following a ‘one-stage approach’,^28^ using a linear mixed-effects (logistic regression) model,^29^ including the trial as a random intercept to account for between-trial variability (see model 1 in **Supplementary Table S3**), and (3) test for HTE through an interaction test by adding the subgroup variable to the mixed-effects model as a main effect and as an interaction term with the treatment variable (see model 3 in **Supplementary Table S3**), and calculated the P value for the interaction term. Additionally, we presented for each subgroup the observed mortality rates in the treatment arms.

To reassess the hypothesized HTE while considering corresponding baseline PEEP settings, we *stratified* patients by observed PEEP tertiles (see section 2.2; step 1). In other words, we repeated steps 1-3 of the reproduction analysis in each PEEP tertile separately. To test whether the HTE between the treatment (ie, lower vs higher PEEP) and PaO_2_/FiO_2_ subgroups varied significantly between the tertiles of baseline PEEP levels (ie, a second order heterogeneity), we performed a second order interaction test. For this, we added the baseline PEEP tertile as main effects and as an interaction term with the treatment variable, and the second order interaction term of treatment, PaO_2_/FiO_2_ subgroup and PEEP tertile to the mixed-effects model (see model 4 in **Supplementary Table S3**). For the first (reproduction) analysis, we only considered patients with non-missing values for baseline PaO_2_/FiO_2_. For the second analysis, we also excluded patients with missing values for baseline PEEP.

#### External validation of ARDS sub-phenotypes

In their 2014 study,^18^ Calfee and colleagues used latent class analysis (LCA) to identify sub-phenotypes in ARDS patients, and found significant HTE in the ALVEOLI trial.^4^ Specifically, patients with the ‘hyperinflammatory’ phenotype benefited from higher PEEP, while those with the ‘hypoinflammatory’ phenotype benefited from lower PEEP in terms of mortality. Since the LCA model relies on lab variables not available in most trials, its use in identifying HTE in other trials is limited. However, Sinha et al.^30^ later developed models using widely available clinical data to classify sub-phenotypes with strong discriminative performance (AUC of 0.95). Although more sparse than the original LCA model, this model still requires a relatively large set of baseline variables, with the LOVS trial^5^ currently being the only available RCT with sufficient data availability. Hence, using the model by Sinha et al.,^30^ we classified all patients (ie, without splitting patients into baseline PEEP tertiles) from the LOVS trial^5^ into hypo- or hyperinflammatory sub-phenotypes and tested for HTE following a one-stage approach (as detailed in the previous section), with the only difference that we will use a fixed-effects (logistic regression) model rather than a mixed-effects model, given that the HTE among the sub-phenotypes will only be evaluated in a single trial (ie, the LOVS trial^5^).

## 3. Preliminary results

### 3.1 Trial and Patient Characteristics

The three trials forming our train cohort consist of 2,299 patients, and the five trials which form our independent (not yet accessed) test cohort consist of 1,404 patients. Consequently, our planned final analyses will include 3,703 patients in the train and test cohorts combined (**Table 1**). IPDs were checked for consistency with the original publications and encountered issues were resolved with the corresponding authors. Inclusion criteria of the different trials were mostly based on the ARDS criteria,^31^ with some variations among the trials. The 28-day mortality rate was notably high in the ART trial^9^ (52.3%), but ranged between 22.6 and 30.4% for seven remaining trials. The PEEP regimes in the lower PEEP (ie, control) arms were similar among the included trials, with seven of the eight trials using the lower PEEP-FiO_2_ table from the ARDSnet protocol,^4^ but varied widely in the experimental arms. For each of the trials, the higher PEEP regimes resulted in higher mean PEEP levels on study day 1 in the experimental groups compared to the control groups (**Supplementary Figure S4a**). The mean FiO_2_ was lower, and the mean PaO_2_/FiO_2_ was higher in the experimental arms compared to the control arms through the first week of the trials (**Supplementary Figures S4b-c**).

**Table 1:** Included trial characteristics. **only for first 80 patients enrolled in the higher PEEP group. RM=recruitment manoeuvre, SRM=Staircase recruitment manoeuvre, PEEP=positive end-expiratory pressure, IPD=individual patient data.

| Reference, year | Cohort | Country | Patients in ITT analysis (n) | Mortality rate (%) | Control strategy |  | Experimental strategy |  |
| --- | --- | --- | --- | --- | --- | --- | --- | --- |
|  |  |  |  |  | RM (PEEP, duration, frequency) | PEEP regime | RM (PEEP, duration, frequency) | PEEP regime |
| **Brower et al., 2004 (ALVEOLI)** | Train cohort (IPD collected) | USA | 549 | 22.6 | None | Lower PEEP-FiO<sub>2</sub> table ARDSnet | 35-40 cmH<sub>2</sub>O PEEP for 30s, 1 or 2 during first 4 days** | Higher PEEP-FiO<sub>2</sub> table (as per Amato et al.<sup>3</sup>), minimum of 12-14 cmH<sub>2</sub>O during first 48h |
| **Meade et al., 2008 (LOVS)** | Train cohort (IPD collected) | Canada | 983 | 30.4 | None | Lower PEEP-FiO<sub>2</sub> table ARDSnet, with plateau pressure < 30 cmH<sub>2</sub>O | 40 cmH<sub>2</sub>O, for 40s, 1 at baseline, and permitted after each ventilator disconnect (up to 4 times/day) until FiO<sub>2</sub> was 40% or less | Higher PEEP table (as per Amato et al.<sup>3</sup>), with plateau pressure < 40 cmH<sub>2</sub>O |
| **Mercat et al., 2008 (EXPRESS)** | Train cohort (IPD collected) | France | 767 | 29.5 | None | As low as possible to meet oxygenation targets (total PEEP 5-9 cmH<sub>2</sub>O) | None | As high as possible while keeping plateau pressure < 28-30 cmH<sub>2</sub>O via incremental PEEP trial |
| **Talmor et al., 2008 (EPVENT)** | Test cohort (IPD not collected yet) | USA | 61 | 27.9 | 40 cmH<sub>2</sub>O, 30s, 1 at baseline | Lower PEEP-FiO<sub>2</sub> table ARDSnet | 40 cmH<sub>2</sub>O, 30s, 1 at baseline | Esophageal-Pressure-Guided, 72 hours |
| **Hodgson et al., 2011 (PHARLAP)** | Test cohort (IPD not collected yet) | Australia | 20 | 25 | None | Lower PEEP-FiO<sub>2</sub> table ARDSnet | Daily staircase RM (SRM) 'SRM' up to 40 cmH<sub>2</sub>O PEEP for 2 min per step, followed by decremental PEEP titration to define 'derecruitment point', followed by brief RM of 40 cmH<sub>2</sub>O for 60s | PEEP 2.5 cmH<sub>2</sub>O above derecruitment point as per daily SRM with decremental PEEP trial (derecruitment point defined by when decreasing PEEP until a decrease in SaO<sub>2</sub> ≥ 1% from maximum SaO<sub>2</sub> was observed, with minimum allowed PEEP 15 cmH<sub>2</sub>O). |
| **Kacmarek et al., 2016** | Test cohort (IPD not collected yet) | USA | 200 | 24.5 | None | Lower PEEP-FiO<sub>2</sub> table ARDSnet | Initial RM with 35-45 cmH<sub>2</sub>O PEEP, for 180s, 1 at baseline | Titrated to PEEP level of highest C<sub>RS</sub> + 2cmH<sub>2</sub>O during decremental PEEP titration, once at baseline |
| **Cavalcanti et al., 2017 (ART)** | Test cohort (IPD not collected yet) | Brazil | 1,010 | 52.3 | None | Lower PEEP-FiO<sub>2</sub> table ARDSnet | Initial SRM up to 45 cmH<sub>2</sub>O (total SRM duration 240s), followed by decremental PEEP trial, followed by second RM of 45 cmH<sub>2</sub>O for 120s<br>Adjustment after 556th patient: SRM up to 35 cmH<sub>2</sub>O (total SRM duration 180s) and second RM of 35 cmH<sub>2</sub>O | PEEP level of highest C<sub>RS</sub> + 2cmH<sub>2</sub>O during decremental PEEP titration |
| **Hodgson et al., 2019 (PHARLAP)** | Test cohort (IPD not collected yet) | Australia | 113 | 25.7 | None | Lower PEEP-FiO<sub>2</sub> table ARDSnet | Initial 'SRM': up to 40 cmH<sub>2</sub>O PEEP for 2 min per step, followed by decremental PEEP titration and brief RM (120s at maximum tolerated PEEP during SRM), daily for 5 days; | PEEP 2.5 cmH<sub>2</sub>O above desaturation level as per daily SRM with decremental PEEP trial (desaturation level defined when decreasing PEEP until SpO<sub>2</sub> decreased |
|  |  |  |  |  |  |  | additional brief RM<br>were allowed<br>throughout the day | by > 2%, with minimum<br>allowed PEEP 15 cmH<sub>2</sub>O). |

RMs were used in both control and experimental arms in one trial,^21^ and only in the experimental arm for six trials, with varying pressure settings and durations of the RM. Pooled baseline characteristics within the train cohort were similar between the control and experimental groups, and also missingness rate of data was similar between treatment arms (**Table 2**).

**Table 2:** Baseline characteristics of the 2,299 patients from the included RCTs in the train cohort. Numbers denote n (%) or median (IQR).

| **Characteristic** | **Higher PEEP**<br>**(N=1,136)** | **Lower PEEP**<br>**(N=1,163)** | **Missingness**<br>**(% higher PEEP,**<br>**% lower PEEP)** |
| --- | --- | --- | --- |
| **Demographics** |  |  |  |
| Female sex, no. (%) | 437 (38.5) | 455 (39.1) | (0.0, 0.0) |
| Age, (years) | 57.3 (43.3-70.0) | 56.0 (43.8-69.7) | (0.0, 0.0) |
| Body Mass Index, (kg/m<sup>2</sup>) | 26.2 (22.9-30.1) | 26.0 (22.6-30.0) | (9.9, 10.5) |
| **Clinical parameters** |  |  |  |
| Resp. rate, (breaths/min) | 22.0 (18.0-28.0) | 23.0 (18.0-28.0) | (0.3, 0.3) |
| Syst. blood pressure, (mmHg) | 115.0 (105.0-128.0) | 115.0 (105.0-128.9) | (0.1, 0.1) |
| Heart rate, (bpm) | 100.0 (85.0-114.0) | 100.0 (85.0-114.0) | (0.1, 0.1) |
| **Blood gasses** |  |  |  |
| PaCO<sub>2</sub>, (mmHg) | 40.0 (35.0-46.0) | 40.0 (35.0-46.0) | (1.0, 1.5) |
| PaO<sub>2</sub>, (mmHg) | 79.0 (67.0-96.0) | 79.0 (68.0-95.0) | (1.0, 1.5) |
| pH | 7.37 (7.32-7.43) | 7.38 (7.32-7.43) | (1.0, 1.5) |
| HCO<sub>3</sub><sup>-</sup>, (mEq/L) | 23.0 (20.0-26.8) | 23.0 (20.0-27.0) | (0.8, 0.9) |
| **Ventilatory parameters** |  |  |  |
| PEEP, (cmH<sub>2</sub>O) | 10.0 (8.0-12.0) | 10.0 (7.0-12.0) | (0.1, 0.3) |
| Tidal volume,<br>(mL/kg predicted body weight) | 7.7 (6.6-9.1) | 7.7 (6.6-9.1) | (2.6, 2.4) |
| PaO<sub>2</sub>/FiO<sub>2</sub>, (mmHg) | 138.3 (103.0-178.0) | 141.7 (101.6-182.5) | (1.0, 1.5) |
| ΔP, (cmH<sub>2</sub>O) | 17.0 (13.0-20.0) | 16.0 (12.0-20.0) | (15.9, 19.1) |
| C<sub>RS</sub>, (mL/cmH<sub>2</sub>O) | 29.4 (22.7-38.0) | 31.2 (24.0-40.0) | (17.7, 21.1) |
| Minute volume, (L/min) | 11.2 (9.2-13.7) | 11.0 (9.1-13.7) | (1.2, 1.0) |
| FiO<sub>2</sub>, (fraction) | 0.6 (0.5-0.8) | 0.6 (0.5-0.8) | (0.0, 0.1) |
| Pplat, (cmH<sub>2</sub>O) | 27.0 (23.0-31.0) | 26.0 (21.0-30.0) | (15.8, 18.7) |

### 3.2 Explorative predictive HTE analysis

#### Step 1: Stratifying patients by set baseline PEEP

Baseline PEEP was missing for four patients, which were consequently excluded from the explorative, predictive HTE analysis. Among the remaining patients, there were 899 with baseline PEEP ≤8 cmH_2_O, 638 with baseline PEEP 9-11 cmH_2_O, and 758 with baseline PEEP ≥12 cmH_2_O (there were no patients with baseline PEEP value between 8 and 9, or between 11 and 12 cmH_2_O).

#### Step 2: Method selection

Among all candidate methods, implemented both with and without forward selection, the IQR of the bootstrapped AUC benefits exceeded zero for only 3 methods (**Figure 4**).

**Figure 4:**
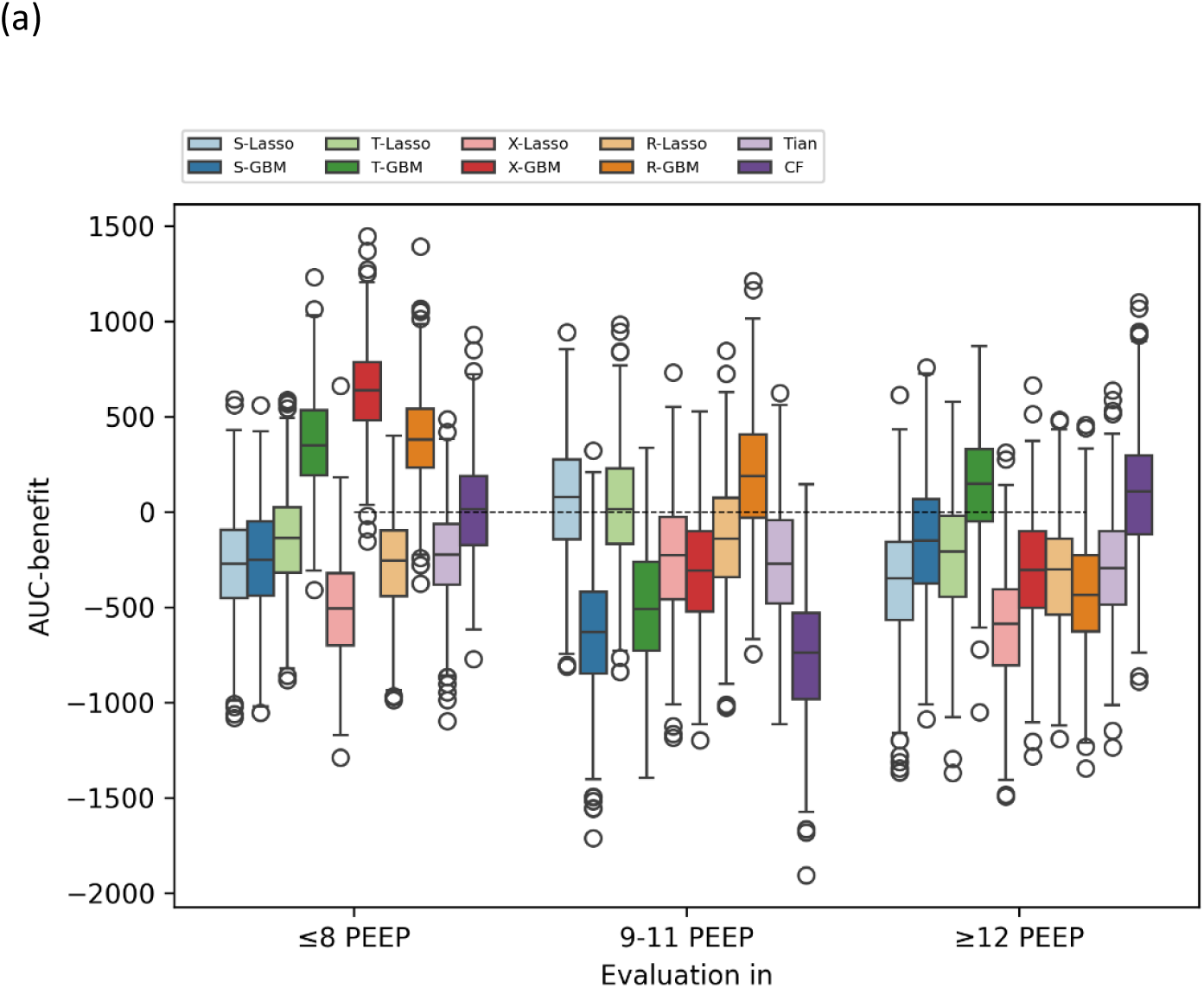
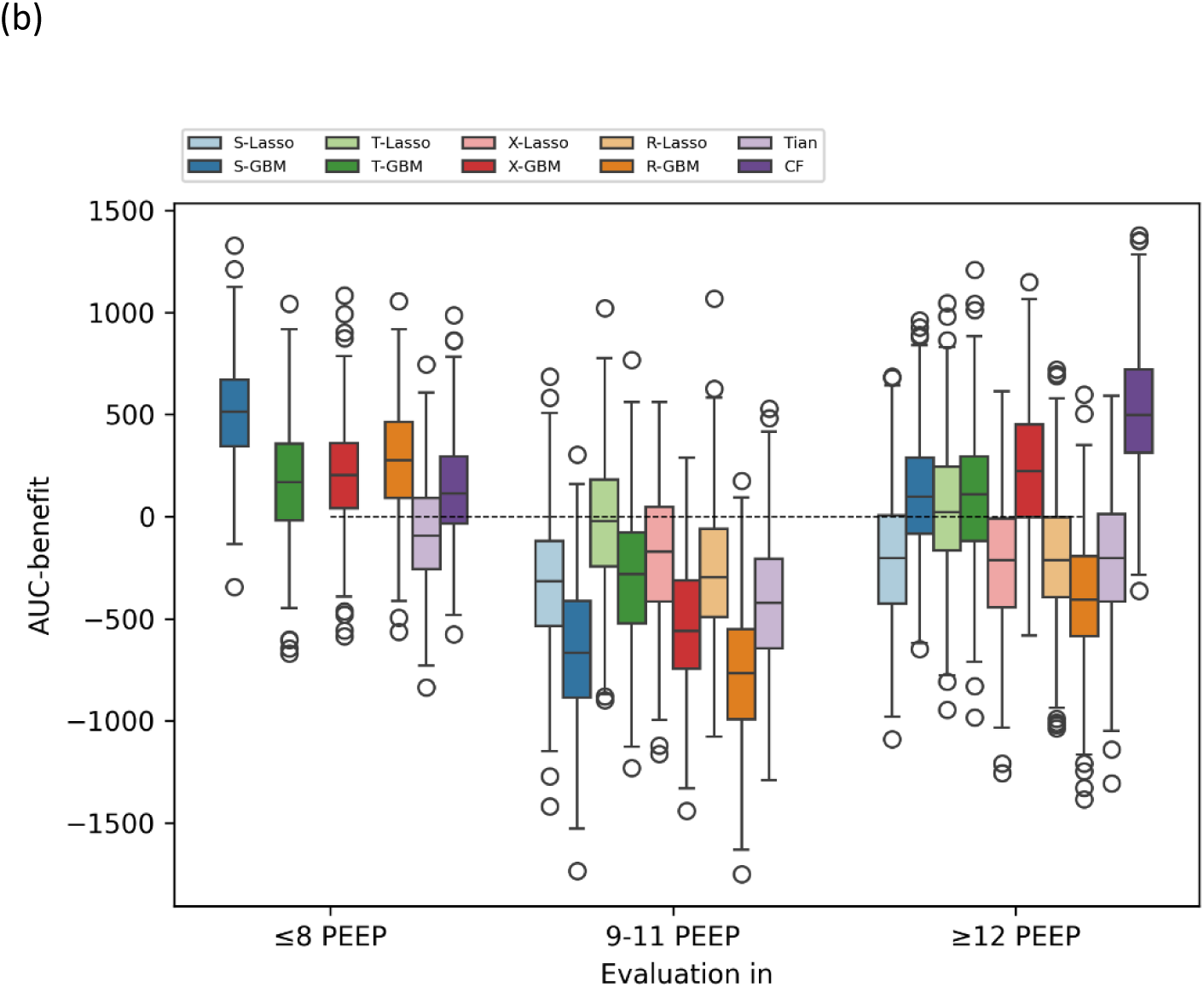
Discrimination for benefit (in terms of AUC-benefit) results of the leave-one-trial-out cross-validation (LOTO-CV) in each of the three PEEP tertiles. Each boxplot represents the AUC-benefits resulting from 500 bootstrap samples. (a) Modelling strategies *excluding* forward selection. (b) Modelling strategies *including* forward selection (for some methods, none of the variables were selected in the forward selection in at least one of the LOTO-CV folds, and therefore the AUC-benefits are not plotted).

In the lower PEEP tertile (PEEP ≤8 cmH_2_O), X-GBM achieved the highest median AUC benefit among methods implemented without forward selection. Among methods implemented with forward selection, the S-GBM achieved the highest median AUC benefit. However, no variables were consistently selected during the forward selections across the outer LOTO-CV folds for S-GBM, precluding the training of an additional final model in this tertile.

In the middle PEEP tertile (PEEP 9-11 cmH_2_O), none of the methods, whether implemented with or without forward selection, demonstrated an AUC-benefit IQR greater than zero.

In the high PEEP tertile (PEEP ≥12 cmH_2_O), none of the methods implemented without forward selection demonstrated an AUC-benefit IQR greater than zero. Among the methods implemented with forward selection, the causal forest method achieved the highest median AUC benefit with an IQR exceeding zero, with C_RS_ being consistently selected during forward selection across all outer LOTO-CV folds (see **Supplementary Table S4**), leading to the training of an additional final model using a causal forest, selecting only C_RS_.

#### Step 3: Training of final models

We trained four final models, using the selected methods using data of the corresponding PEEP tertile of the complete train cohort. In the lower PEEP tertile, we trained a final X-GBM model without forward selection (ie, ‘final model 1’), and an S-GBM with forward selection (ie, ‘final model 2’). Final model 2 selected two variables, ie, driving pressure and the binary variable of pulmonary ARDS (yes/no; see **Supplementary Table S5a**). In the high PEEP tertile, we trained a causal forest with forward selection (ie, ‘final model 3’), which selected two variables, ie, driving pressure and tidal volume (mL/kg predicted body weight; see **Supplementary Table S5b**), and trained a causal forest selecting only C_RS_ as a feature (ie, ‘final model 4’). Because final model 4 consists of only one variable (in contrast to the other trained final models), each predicted individualized treatment effect corresponds to a certain C_RS_ value. **Figure 5** shows the predictions corresponding to a wide range of C_RS_ values, showing that the predictions turn from negative (ie, predicted harm from high PEEP) to positive (ie, predicted benefit from high PEEP) around a C_RS_ value of 26.5 mL/cmH_2_O. Notably, the individualized treatment effect predictions display a ‘U-shaped’ curve, shifting from harm to benefit after 26.5 mL/cmH_2_O, then decreasing again at C_RS_ values around 40 mL/cmH_2_O and above. However, due to the scarcity of patients with C_RS_ ≥ 60 mL/cmH_2_O in the training cohort, the model lacks sufficient data to predict the treatment effect of higher PEEP in this range. Consequently, patients with baseline C_RS_ > 60 mL/cmH_2_O might also experience harm from higher PEEP, similar to those with C_RS_ < 26.5 mL/cmH_2_O, but this cannot be predicted by the model.

**Figure 5:**
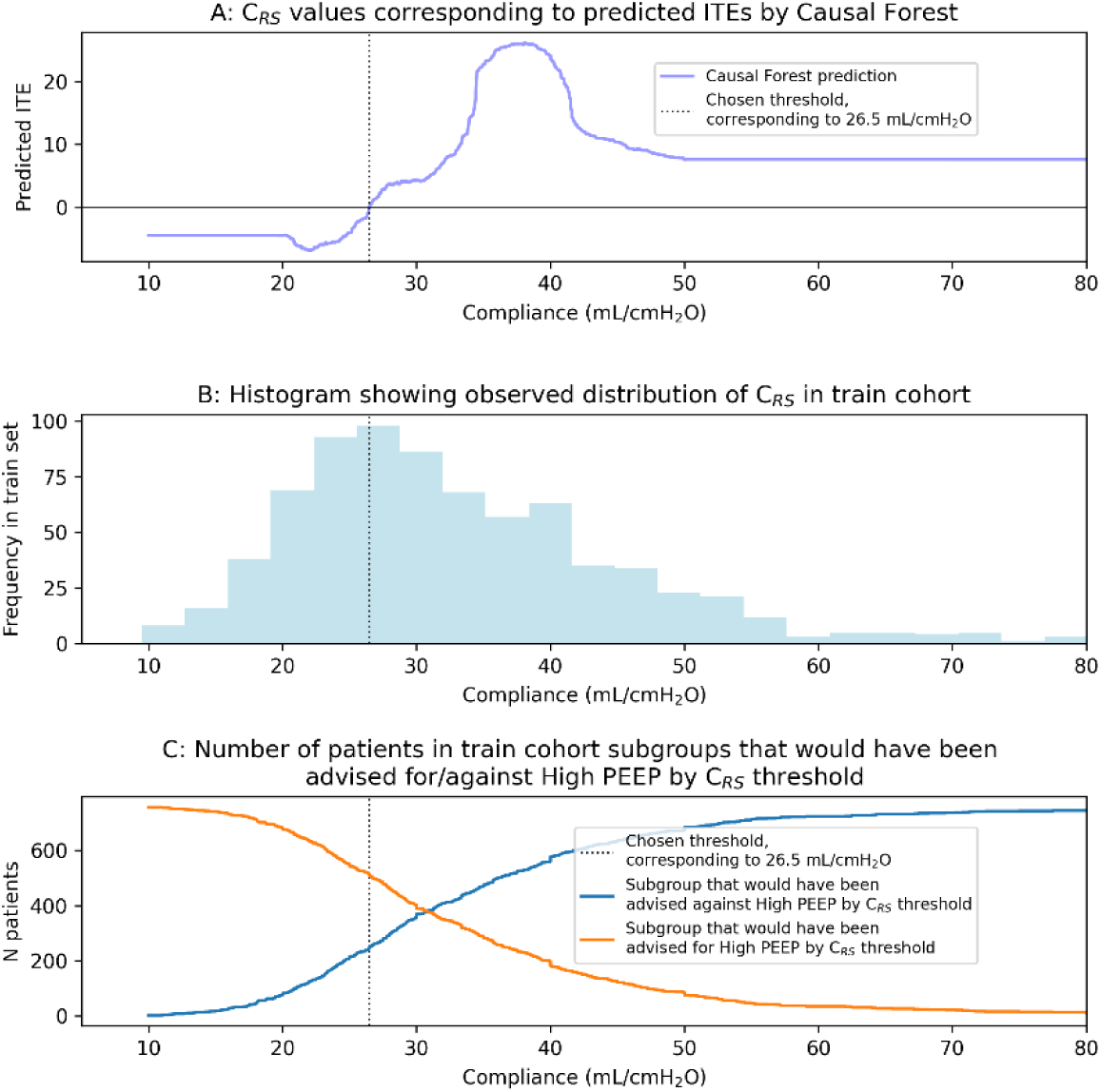
Because C_RS_ is the only variable in final model 4, each individualized treatment effects (ITEs) predicted by this model corresponds to a C_RS_ value. (A) predicted ITEs corresponding to each possible C_RS_ value between 10 and 80 cmH_2_O. (B) The observed C_RS_ distribution in the train cohort. (C) The sizes of the patient subgroups the would result from different decision thresholds (ie, the ITE value above which treating patients is considered worthwhile).

#### Step 4: Pre-registering of final models

All the trained final models, as well as the corresponding dependencies (ie, trained scalers and imputers), can be found on Github.^32^

### 3.3 Evaluation of earlier hypothesized HTE

#### A closer look at Briel’s hypothesis

Consistent with Briel et al.’s^7^ findings, higher PEEP improved outcomes for patients with a baseline PaO_2_/FiO_2_ ≤ 200 mmHg, with 28-day mortality rates of 33% in the lower PEEP group versus 28% in the higher PEEP group (OR 0.80 [95% CI 0.66 to 0.98]). Conversely, for patients with baseline PaO_2_/FiO_2_ > 200 mmHg, higher PEEP was associated with worse outcomes, with mortality rates of 14% in the lower PEEP group compared to 22% in the higher PEEP group (OR 1.74 [95% CI 1.02 to 2.98]). This HTE was statistically significant (P for interaction = 0.01; **Figure 6**).

**Figure 6:**
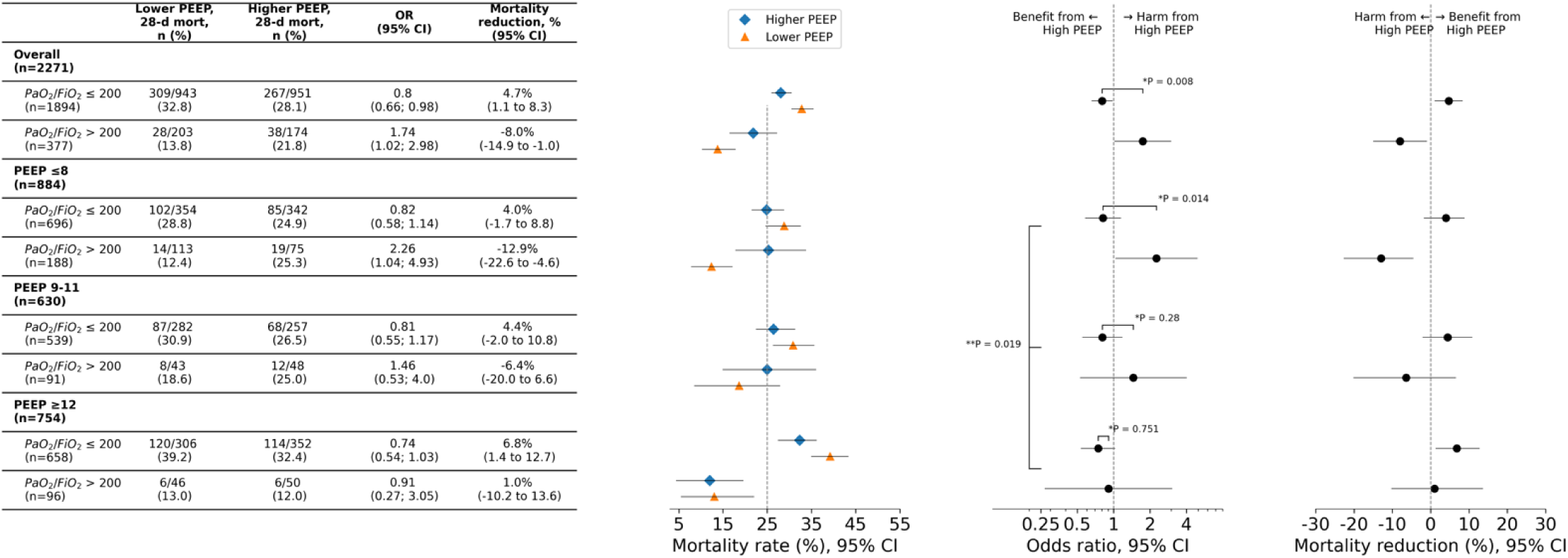
Treatment effects of higher vs lower PEEP on the relative, odds ratio scale and the absolute, mortality risk difference scale, plotted for patients with baseline PaO_2_/FiO_2_ > 200 mmHg and baseline PaO_2_/FiO_2_ ≤ 200 mmHg, for the full train cohort (‘Overall’) and separately for baseline PEEP tertiles. *P value for interaction between PaO_2_/FiO_2_ subgroup and treatment, ie, first-order interaction. **P value between PEEP tertile, PaO_2_/FiO_2_ subgroup and treatment, ie, second-order interaction.

Notably, the HTE between PaO_2_/FiO_2_ subgroups was most pronounced when PaO_2_/FiO_2_ was measured at low baseline PEEP (≤ 8 cmH2O), diminished at middle PEEP (9–11 cmH2O), and further reduced at higher PEEP (≥ 12 cmH2O). At low PEEP, mortality for PaO_2_/FiO_2_ ≤ 200 mmHg was 29% in the lower PEEP group and 25% in the higher PEEP group (OR 0.82 [95% CI 0.58 to 1.14]), while for PaO_2_/FiO_2_ > 200 mmHg, mortality was 12% versus 25% (OR 2.26 [95% CI 1.04 to 4.93]; P for interaction = 0.01). At middle PEEP, patients with PaO_2_/FiO_2_ ≤ 200 mmHg had mortality rates of 31% in the lower PEEP group versus 27% in the higher PEEP group (OR 0.81 [95% CI 0.55 to 1.17]), while for PaO_2_/FiO_2_ > 200 mmHg, mortality was 19% versus 25% (OR 1.46 [95% CI 0.53 to 4.00]; P for interaction = 0.28). At high PEEP, mortality for PaO_2_/FiO_2_ ≤ 200 mmHg was 39% in the lower PEEP group versus 32% in the higher PEEP group (OR 0.74 [95% CI 0.54 to 1.03]), while for PaO_2_/FiO_2_ > 200 mmHg, it was 13% versus 12% (OR 0.91 [95% CI 0.27 to 3.05]; P for interaction = 0.75).

Hence, the treatment effect of higher versus lower PEEP showed the greatest heterogeneity between baseline PaO_2_/FiO_2_ subgroups when PaO_2_/FiO_2_ was measured at low PEEP. A second-order interaction test confirmed that this HTE varied significantly across baseline PEEP tertiles (P-value for second-order interaction = 0.03; **Figure 6**).

#### External validation of ARDS sub-phenotypes

The model by Sinha et al.,^30^ using readily available clinical data, classified 843/983 (85.8%) of the patients in the LOVS trial^5^ to the hypoinflammatory, and 140/983 (14.2%) to the hyperinflammatory sub-phenotype. In line with findings from the ALVEOLI trial regarding these subphenotypes,^18^ the hyperinflammatory subphenotype exhibited a higher 28-day mortality rate compared to the hypoinflammatory subphenotype (50.7% versus 27.0%). For patients in the hypoinflammatory phenotype, we observed 28-day mortality rates of 29.3% in the lower PEEP group versus 24.6% in the higher PEEP group (OR 0.79 [95% CI 0.58 to 1.07]). For patients in the hyperinflammatory phenotype, we observed 28-day mortality rates of 53.1% in the lower PEEP group versus 48.7% in the higher PEEP group (OR 0.84 [95% CI 0.43 to 1.63]). Hence, the treatment effect of higher PEEP was not significantly heterogeneous between the predicted subphenotypes (P for interaction = 0.512; **Figure 7**).

**Figure 7:**
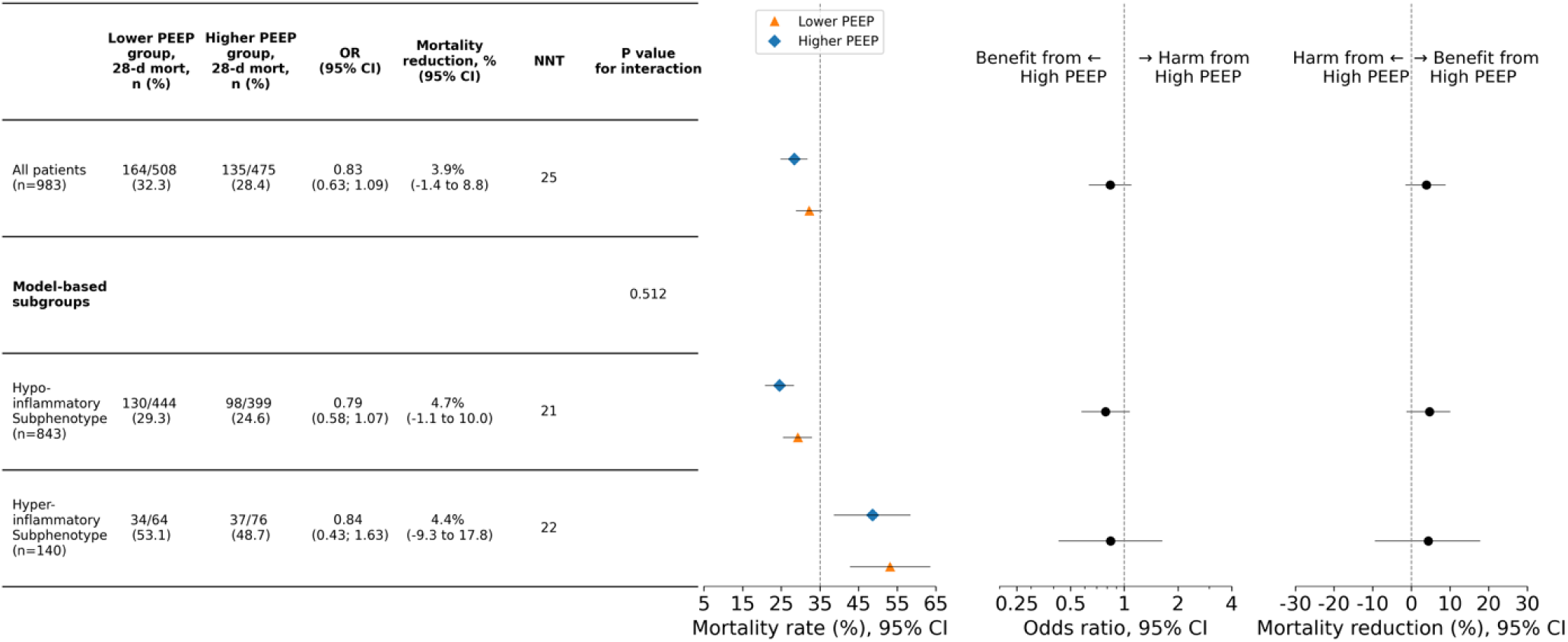
Treatment effects of higher vs lower PEEP on the relative, odds ratio scale and the absolute, mortality risk difference scale, plotted for patients from the LOVS trial^5^, classified into the hypo- and hyperinflammatory phenotype, as predicted by the model developed by Sinha et al.^30^ NNT=number needed to treat.

## 4. Protocol for external validation

### 4.1 Explorative Predictive HTE analysis

#### Data pre-processing

A scaler and imputer will first be applied to normalize the data from the test cohort and subsequently impute missing values. Since the final models were trained within the same PEEP tertile using the same a priori selected variables (see **Supplementary Table S1**), the scalers (i.e., variable means and standard deviations used for normalization) and trained imputers were identical for models trained within each PEEP tertile. Hence, we trained and saved a scaler and imputer using data from the train cohort in the lower PEEP tertile for pre-processing data in final models 1 and 2. Similarly, a scaler and imputer were trained and saved using data from the train cohort in the higher PEEP tertile for pre-processing data in final models 3 and 4.

#### Evaluation of HTE

For each of the final models, we will predict individualized treatment effects for the patients in the test cohort for whom the variables in the model were measured during a PEEP level corresponding to the PEEP tertile in which the model was trained (**Figure 2**). The resulting individualized treatment effect predictions will be evaluated in terms of discrimination for benefit, calibration for benefit, and the statistical significance of the HTE between the patient subgroups resulting from these predictions. Each component of the evaluation will be performed in the ‘merged’ test cohort, in which all the patients from the trials are handled as coming from one large trial, as well as in each of the trials in the test cohort separately. Given the small size of the Hodgson 2011 trial, the patients from the Hodgson 2011 and Hodgson 2019 trials will always be merged.

- *Discrimination for benefit:*

We will assess discrimination for benefit for each final model in the test cohort, calculating the weighted average of the AUC-benefits resulting from the individual trials in the test cohort.

- *Calibration for benefit:*

We will evaluate the calibration for benefit by dividing patients into four groups based on ascending predicted individualized treatment effect quartiles and plotting these predictions alongside the observed mortality reductions.

- *Test for statistical significance of HTE:*

To test for HTE among the subgroups identified by these models, we will first assume a decision threshold (ie, the value above which treating patients is considered worthwhile^33^) of ITE=0, dividing patients into a subgroup for whom the model predicted benefit from high PEEP (ie, ITE > 0) and a subgroup for whom the model predicted harm from high PEEP (ITE ≤ 0; **Figure 1**). For the resulting subgroups, we will estimate relative treatment effects in terms of marginal odds ratios (ORs) in a one-stage approach^28^ using a linear mixed-effects model (logistic regression), including the trial as a random intercept to account for between-trial variability (see model 1 in **Supplementary Table S3**). We will test for HTE through an interaction test by adding the subgroup variables to the mixed-effects model as a main effect and as an interaction term with the treatment variable (see model 3 in **Supplementary Table S3**), and calculate the P value for the interaction term.

- *Reporting of treatment effect measures:*

Further, following the reporting guidelines by Kent et al.,^34^ we will report observed mortality rates in each of the treatment arms (ie, the lower and the higher PEEP strategy), relative and absolute treatment effects in terms of odds ratios and mortality rate reductions, respectively, and numbers needed to treat (NNTs) among the subgroups identified by both models (ie, patients with ITE ≤ 0 vs ITE > 0) in the test cohort, the train cohort, and in the full cohort (ie, all eight trials in the train and test cohorts combined).

#### Overall effect analysis

To evaluate the *overall* effect of higher vs lower PEEP strategies on the primary endpoint, ie, in all ARDS patients regardless of subgroups, we will estimate relative treatment effects using the same one-stage approach^28^ using a linear mixed-effects model (logistic regression), including the trial as a random intercept to account for between-trial variability (see model 1 in **Supplementary Table S3** for the exact Python implementation), and report observed mortality rates, treatment effects, and NNTs based on the full cohort (ie, all eight included trials). Additionally, we will assess the overall effects of higher vs lower PEEP strategies in the train and test cohort separately, and in each individual trial.

### 4.2 Sensitivity analyses

#### Exclusion of ART trial patients with initial RM strategy

While the ART trial^9^ was being conducted, starting with the 556th patient, the steering committee decided to modify the RM and PEEP titration strategy after 3 cases of resuscitated cardiac arrest possibly associated with the experimental group treatment were observed. As the initial RM and PEEP titration strategy randomized in this trial are potentially related to harmful effects, we will conduct a sensitivity analysis, repeating the evaluation of HTE (as described in 5.2.1, Main analysis), but excluding the patients from the test cohort who were among the first 555 enrolled patients in the ART trial.

#### Exclusion of trial patients with ‘staircase’ RM strategies

Although all trials in that form the test cohort performed an RM in the higher PEEP treatment arm (with the trial by Talmor et al.^21^ being the only one with RMs in both the lower and high PEEP arms), in three of these trials, ie, Hodson et al 2011^10^ and 2019^11^ and Cavalcanti et al.^9^, a so-called ‘staircase’ RM (SRM) was performed (**Table 1**). An SRM involves incrementally increasing airway pressure in steps to recruit collapsed alveoli, followed by a gradual decrease to identify the optimal pressure that maintains lung recruitment while minimizing overdistension. As SRMs might be related to harmful effects, we will conduct a sensitivity analysis, repeating the evaluation of HTE (as described in 5.2.1, Main analysis), but excluding the patients from the test cohort from the three trials^9–11^ with SRMs, hence only using patients from the trials by Talmor et al.^21^ and Kacmarek et al.^8^

#### Risk modelling

Following the HTE framework as proposed by Kent and colleagues,^17^ who categorize predictive HTE approaches into risk and effect modelling, we will benchmark our proposed effect modelling approach with risk modelling. Each of the included trials has collected a baseline risk score for the included patients using a wide-spread and validated, multivariate disease severity model. Among the eight eligible trials, five^5,8,10,11,21^ collected APACHE II scores, two^4,10^ collected APACHE III scores, one^6^ collected SAPS II scores and one^9^ collected SAPS III scores. We will stratify patients into risk groups based on observed quartiles of these scores, and test for HTE by the respective severity scores by performing interaction tests between treatment effect and the scores, and calculate the P-value of the interaction term (see model 6 in **Supplementary Table S3**).

#### Conditional effects

Addressing the non-collapsibility of the marginal OR measure, we will also calculate ORs conditional on three strong and widely available prognostic factors for the primary endpoint, which we will identify by fitting a logistic regression model on the complete cohort (ie, the train and test cohort combined), and select the three variables with the largest absolute weights. For each calculated OR, we will subsequently calculate conditional ORs through ‘direct adjustment’ (see model 2 in **Supplementary Table S3**).

#### Aggregation bias

We will adopt the one-stage approach^28^ to test the significance of the HTE (see 4.1, test for significance of HTE). This allows between-trial information to contribute toward the summary interaction estimate (which represents the HTE), in combination with within-trial information, which may lead to aggregation bias (also known as ecological bias).^28^ In other words, aggregation bias occurs when relationships observed between trials, such as a variable-treatment interaction, do not hold at the individual patient level, leading to incorrect conclusions about individuals based on group data. Riley and colleagues^28^ proposed a method to disentangle within-trial and between-study information in the one-stage model by centring the subgroup covariate about the trial-specific means, and including the trial-specific mean as an additional adjustment term to explain between-trial heterogeneity (see model 5 in **Supplementary Table S3**). However, as the test cohort includes only five trials, among which three consist of very few patients (<100), this will limit accurately modelling between-trial heterogeneity and, therefore, the reliability of the adjustment method.

Therefore, in addition to this adjustment method, we will examine the HTE within each of the eight individual trials. Although this approach cannot test the full absence of aggregation bias, if HTE in the same direction is observed in most of the individual trials, this will be an indication that any observed HTE in the test cohort is primarily driven by within rather than between-trial effects.

#### Bias due to missing outcomes

In case the mortality outcome is missing for certain patients in the test cohort (eg, due to loss to follow-up), we will exclude these patients from the main analysis. If this missingness of the outcome data is related to prognostic factors at baseline as well as treatment group, exclusion of these patients will create a baseline imbalance in prognosis potentially leading to biased effect estimates.^35^

Therefore, in case of missingness of the mortality outcome variable in the test cohort, we will examine the relationship of missing outcome with treatment group (by comparing the missingness rates among the two treatment arms) and baseline prognostic factors (by comparing distributions of prognostic factors for among patients with missing and non-missing outcomes).

#### Complete case analysis

As we pre-register to impute missing values of variables needed in the final models (section 4.1; pre-processing), we will examine the effect of imputation by repeating the evaluation of the HTE (section 4.1; test for statistical significance of HTE) for each final model, only using data of patients with non-missing values for the variables included in each of the final models (ie, complete case analysis).

#### Examine robustness of Imputation method

As we pre-register only one imputation method (ie, KNN-imputation with K neighbours = 5), we will examine the robustness of our findings for the chosen imputation method by repeating the evaluation of the HTE for each final model while varying the ‘K’ parameter of the KNN imputer between 3 and 20. Additionally, we will repeat the evaluation of the HTE for each final model using an alternative imputation method, scikit-learn’s ‘IterativeImputer’. This imputation method (inspired by R’s MICE package) imputes each variable with missing values based on the remaining variables with Bayesian ridge regression in an iterated round-robin fashion.

### 4.3 Re-assessment of earlier hypothesized HTE

#### A closer look at Briel’s hypothesis

After publication of Briel et al.’s IPDMA In 2010,^7^ all published and upcoming RCTs^8–13^ excluded patients with baseline PaO_2_/FiO_2_ > 200 mmHg. Hence, our hypothesis (based on the train cohort) that treatment effect is most heterogeneous between patients with baseline PaO_2_/FiO_2_ ≤ 200 mmHg vs PaO_2_/FiO_2_ > 200 mmHg when this PaO_2_/FiO_2_ was measured at low PEEP, cannot be further validated in these trials. Apart from the trials in the train cohort, only the trial by Talmor et al.^21^ did not exclude patients with baseline PaO_2_/FiO_2_ > 200 mmHg, and could therefore still be used to further validate this finding. As this trial is relatively small (ie, 61 patients in the intention-to-treat analysis), it is unlikely to observe statistically significant HTE comparing patients with baseline PaO_2_/FiO_2_ ≤ 200 mmHg vs PaO_2_/FiO_2_ > 200 mmHg in this trial alone, although the direction of the HTE may still be informative for the robustness of this hypothesis.

Additionally, as the remaining trials in the test cohort did include patients with PaO_2_/FiO_2_ ≤ 200, measured at varying PEEP settings, these data could still reduce the variance of the effect measures in the patient subgroups with PaO_2_/FiO_2_ ≤ 200, also leading to lower variance for the second-order interaction test. Hence, we will also repeat the evaluation of Briel’s hypothesis, as described in section 2.3.1, using data from all the eight trials (ie, the train and test cohorts combined).

## 5. Apparent validation of final models

Using the same data used to train the final models (ie, ‘apparent’ validation^36^), we evaluated the HTE resulting from the final models in the same fashion as we will evaluate the final models in the external validation (section 4.1; *Evaluation of HTE*). Hence, we used final models 1-4 to predict individualized treatment effects for the patients in the PEEP tertiles based on which each model was trained (ie, only in patients with baseline PEEP ≤ 8 cmH_2_O for final models 1 and 2, and only patients with baseline PEEP ≥12 cmH_2_O for final models 3 and 4.

The treatment effect measures among subgroups identified by the final models, as well as results of the tests for statistical significance of HTE, are given in **Figure 8**. For all final models, we found statistically significant HTE (ie, P-value for interaction <0.01) between the subgroups with predicted harm, and predicted benefit from high PEEP. For each of the final models, the HTE was found consistently in the three individual trials which comprise the train cohort (ie, ALVEOLI^4^, LOVS^5^ and EXPRESS^6^; **Supplementary Figures S5-8**). **Figure 9** depicts the results of the apparent validation in terms of calibration for benefit, including the corresponding AUC-benefits. Notably, the calibration plots final models 1-3 (**Figure 9 a-c**) show reveal too moderate individualised treatment effect predictions, with more severe harm observed than predicted in the lower individualised treatment effect region, and more benefit observed than predicted in the higher individualised treatment effect region. Final model 4 yielded a better calibration for benefit, with predicted individualised treatment effects more closely matching the observed treatment effects in each region (**Figure 9d**).

**Figure 8:**
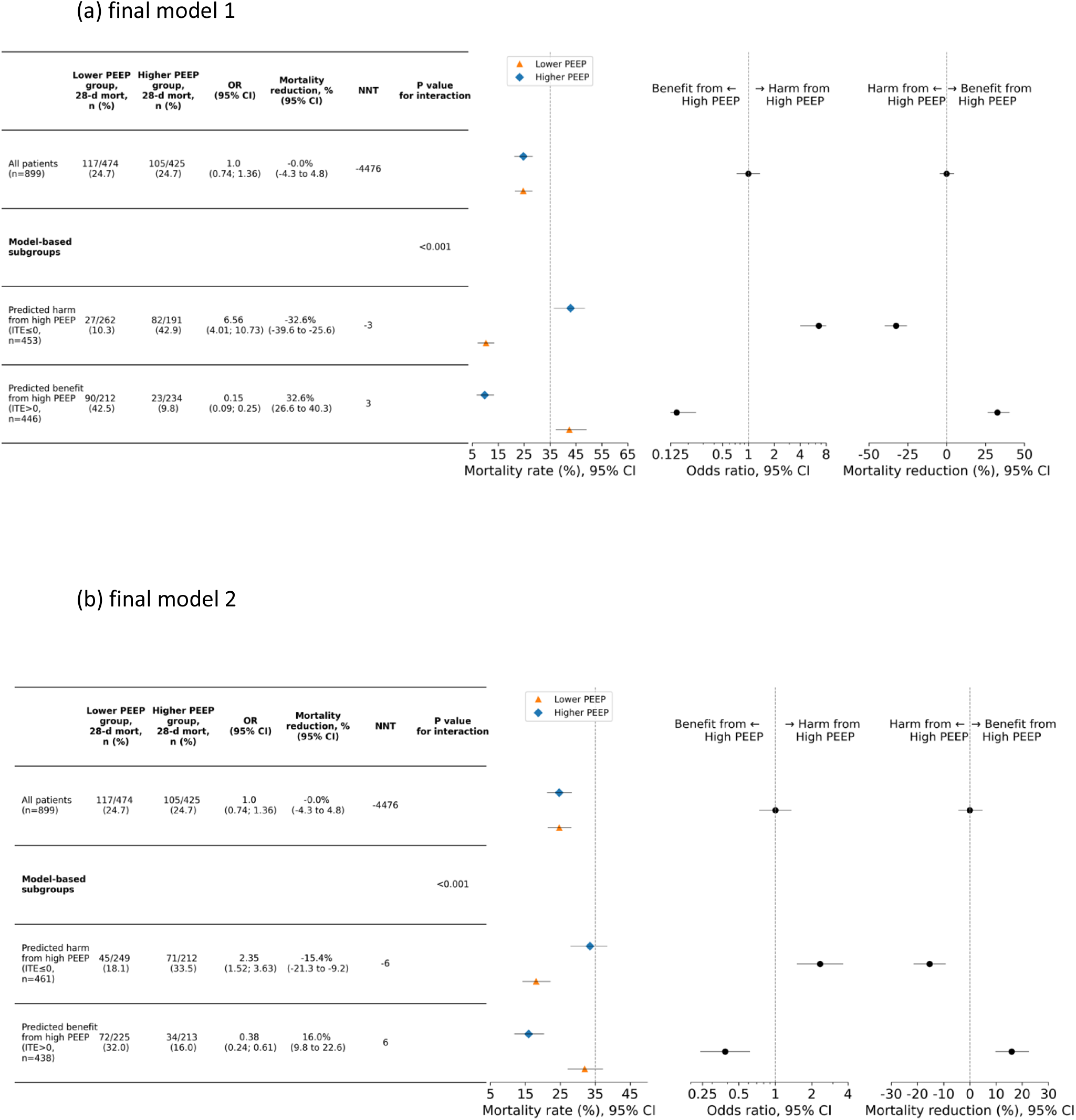
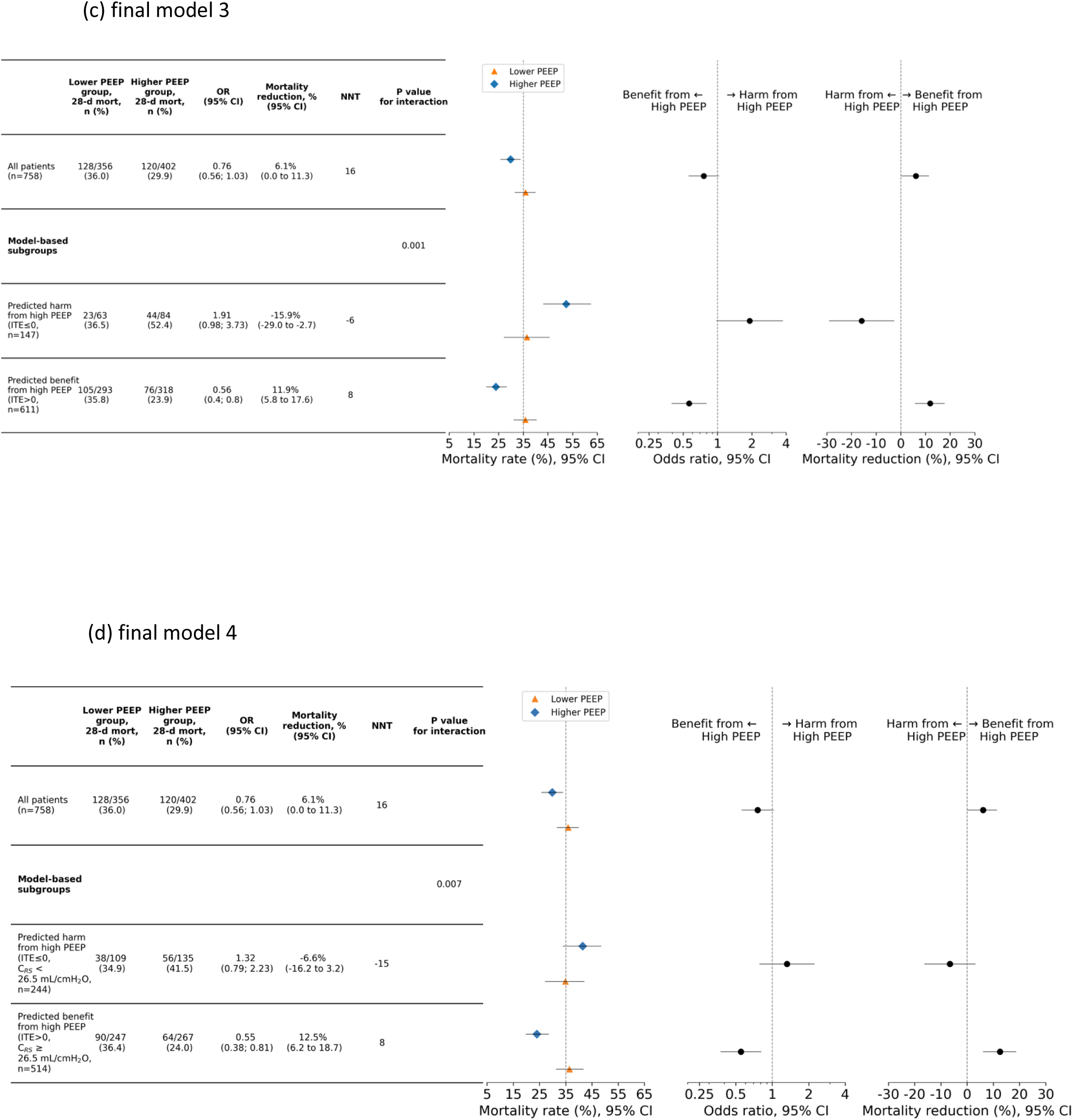
Heterogeneity of treatment effect results of ‘apparent’ validation (ie, models both trained and evaluated in train cohort). Treatment effects of higher vs lower PEEP on the relative, odds ratio scale and the absolute, mortality risk difference scale, (a-b) plotted for patients from the train cohort with baseline PEEP ≤ 8 cmH_2_O (n=899), classified into the predicted harm, and predicted benefit from high PEEP by final models 1 and 2, and (c-d) patients from the train cohort with baseline PEEP ≥12 cmH_2_O (n=758), classified into the predicted harm, and predicted benefit from high PEEP by final models 3 and 4. NNT=number needed to treat.

**Figure 9:**
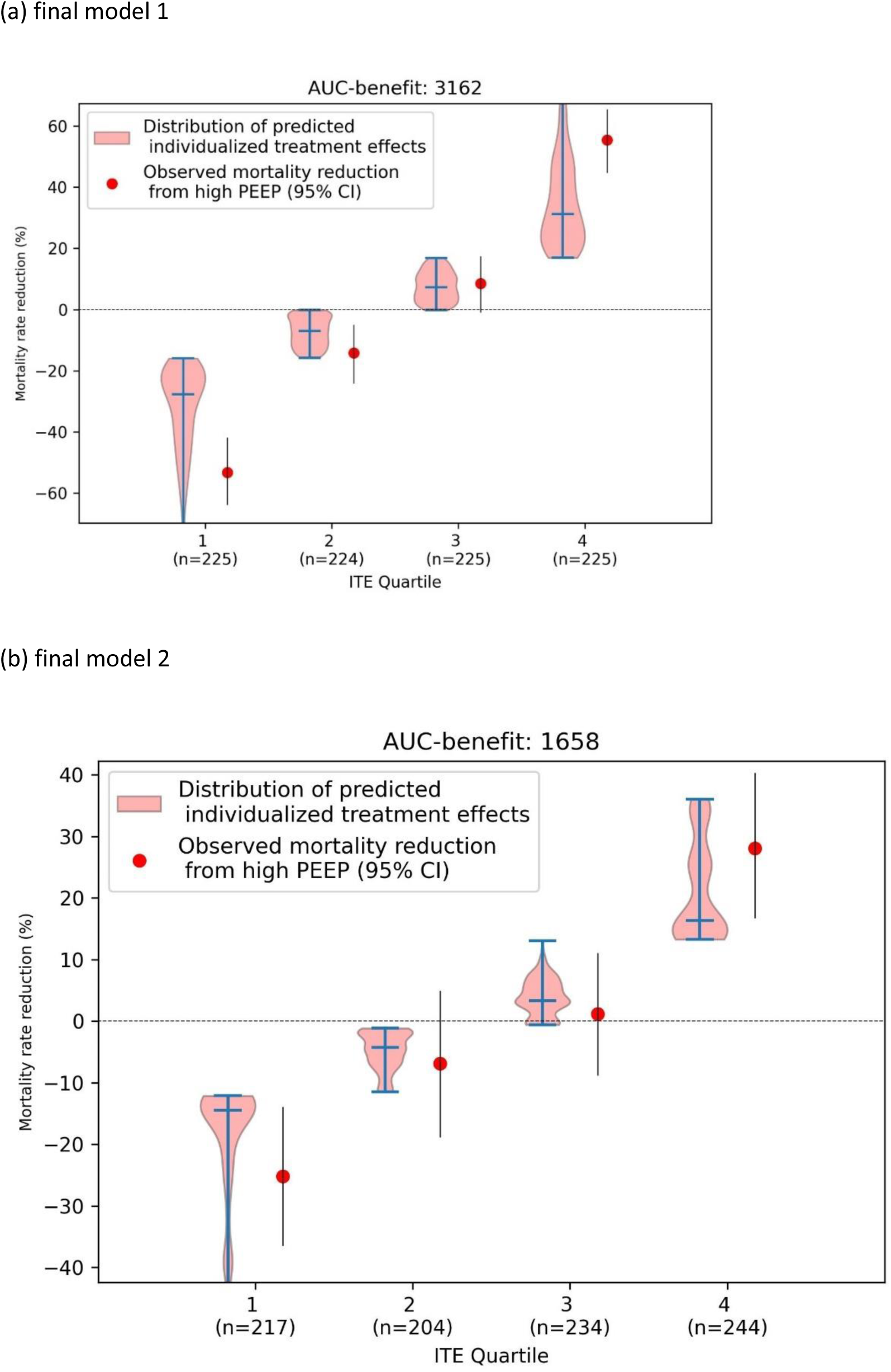
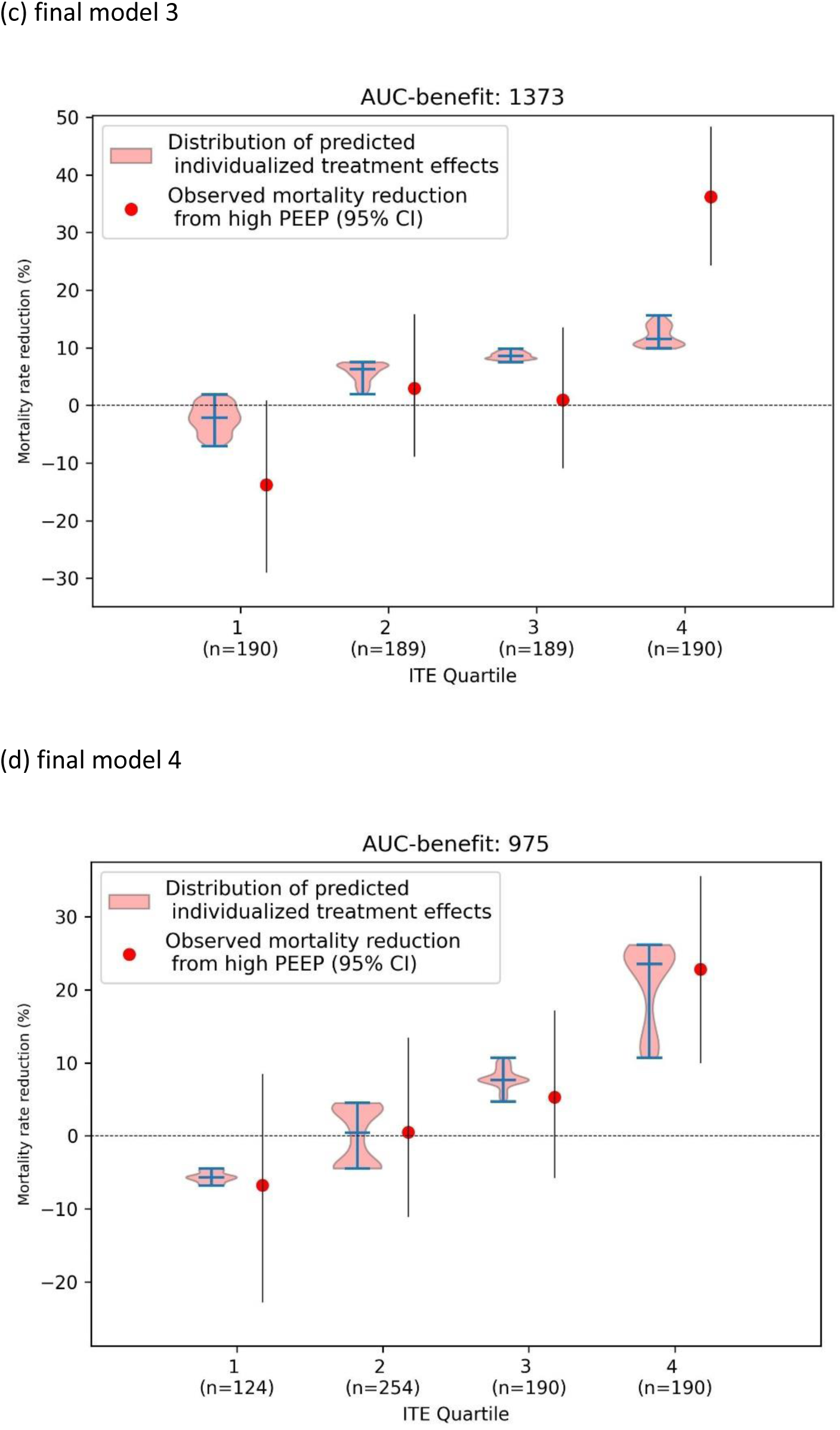
Calibration for benefit results, including AUC-benefit, of ‘apparent’ validation (ie, models both trained and evaluated in train cohort). For patients in the train cohort (a-b) with baseline PEEP ≤ 8 cmH_2_O (n=899) and (c-d) with baseline PEEP ≥12 cmH_2_O (n=758), patients are split into four subgroups based on ascending quartiles of the predicted individualised treatment effects (ITEs) predicted by the corresponding final models. The predicted ITE distributions are visualised using violin plots next to the observed mortality reductions in each quartile. Error bars indicate 95% CIs.

**Supplementary Figures S9-12** depict the results of the apparent validation in terms of calibration for benefit, including the corresponding AUC-benefits, in the three individual trials which comprise the train cohort (ie, ALVEOLI^4^, LOVS^5^ and EXPRESS^6^), again suggesting better calibration by final model 4 in each of the individual trials.

## 6. Preliminary conclusions

We trained four final models (available on Github^32^) using our train cohort, consisting of the ALVEOLI^4^, LOVS^5^ and EXPRESS^6^ trial. Using patients form the train cohort with baseline PEEP ≤8 cmH_2_O, we fitted an X-GBM model without forward selection (‘final model 1’) and an S-GBM model based on driving pressure and pulmonary ARDS (yes/no) selected by forward selection (final model 2). Using patients form the train cohort with baseline PEEP ≥12 cmH_2_O, we fitted a causal forest based on driving pressure and tidal volume selected by forward selection (final model 3) and a causal forest based only on C_RS_ (final model 4). Final model 4 indicates that baseline C_RS_ ≥ 26.5 mL/cmH_2_O predicts benefit, while C_RS_ < 26.5 mL/cmH_2_O predicts harm from high PEEP when C_RS_ is measured at a high baseline PEEP (≥12 cmH_2_O). Each of the four final models will be externally validated using the - yet to be shared-data of the five trials^8–11,21^ forming the test cohort.

Our train cohort includes trials with varying recruitment maneuver (RM) protocols: some applied RMs in the higher PEEP arm for a subset of patients (ALVEOLI), others applied RMs to all patients in the treatment arm (LOVS), while some did not randomize RMs at all (EXPRESS). By merging these datasets as if from one large trial, our modeling strategy implicitly assumes the ability to identify patients who benefit from high PEEP, regardless of RM use.

Based on the train cohort, we revisited earlier hypothesized HTE from high PEEP between patient subgroups based on baseline PaO_2_/FiO_2_. We confirmed this result, but also found that this heterogeneity itself is heterogeneous among baseline PEEP levels during which the baseline PaO_2_/FiO_2_ was measured. According to this new finding, baseline PaO_2_/FiO_2_ ≤ 200 mmHg predicts benefit, while PaO_2_/FiO_2_ > 200 mmHg predicts harm from high PEEP, when PaO_2_/FiO_2_ is measured at a low baseline PEEP (≤8 cmH_2_O).

Using data from the LOVS trial^5^, we examined HTE for high PEEP between patients classified as hypo- and hyperinflammatory subphenotype, but did not observe any HTE, although significant HTE was observed earlier between these subphenotypes in the ALVEOLI trial.^18^

## Data Availability

All data produced in the present study are available upon reasonable request to the authors

