## Supplementary material for "The Heterogeneous Effect of High PEEP strategies on Survival in Acute Respiratory Distress Syndrome: preliminary results of a data-driven analysis of randomized trials"

**Corresponding author:**

### Outline

**Appendix part 1:** Supplementary Figures and Tables

**Appendix part 2:** Definition of the ‘Area under the benefit curve’ (AUC-benefit)

**Appendix part 1:** Supplementary Figures and Tables

Supplementary Table S1: Lists of the a-priori selected variables based on availability in the train and test cohorts, for each of the three PEEP tertiles.

| | **A-priori selected variable set** |
|---|---|
| ***Lower PEEP tertile ($\leq$8 $cmH_2O$)*** ***&*** ***Higher PEEP tertile ($\geq$12 $cmH_2O$)*** | • Sex (0=Female, 1=Male)<br>• Pulmonary ARDS (0=no, 1=yes)<br>• Age (years)<br>• Heart rate (bpm)<br>• Minute Volume (L/min)<br>• Plateau Pressure ($cmH_2O$)<br>• $FiO_2$ (%)<br>• Tidal Volume (mL/kg predicted body weight)<br>• Respiratory Rate (breaths per minute)<br>• Driving Pressure ($cmH_2O$)<br>• Respiratory System Compliance ($mL/cmH_2O$) |
| ***Middle PEEP tertile (9-11 $cmH_2O$)*** | • Sex (0=Female, 1=Male)<br>• Pulmonary ARDS (0=no, 1=yes)<br>• Age (years)<br>• Heart rate (bpm)<br>• Minute Volume (L/min)<br>• Plateau Pressure ($cmH_2O$)<br>• $FiO_2$ (fraction)<br>• Tidal Volume (mL/kg predicted body weight)<br>• Respiratory Rate (breaths per minute)<br>• Driving Pressure ($cmH_2O$)<br>• Respiratory System Compliance ($mL/cmH_2O$)<br>• $PaO_2$ (mmHg)<br>• $PaCO_2$ (mmHg)<br>• pH<br>• $PaO_2/FiO_2$ (mmHg) |

Supplementary Table S2: Searched grids of the hyperparameters of the 10 candidate methods.

| ***Effect modelling technique*** | **Hyperparameter** | **Grid searched** |
|---|---|---|
| *S-Lasso* | penalty strength | $10^{-2}$ to $10^{2}$, evenly spaced on a logarithmic scale (7 steps) |
| *S-GBM* | Boosting type | ['gbdt', 'dart'] |
| | Maximum tree depth | |
| *T-Lasso* | **Outcome model in control group:** penalty strength | $10^{-2}$ to $10^{2}$, evenly spaced on a logarithmic scale (7 steps) |
| | **Outcome model in treatment group:** penalty strength | $10^{-2}$ to $10^{2}$, evenly spaced on a logarithmic scale (7 steps) |
| *T-GBM* | **Outcome model in control group:** Boosting type | ['gbdt', 'dart'] |
| | **Outcome model in control group:** Maximum tree depth | [3, unlimited] |
| | **Outcome model in treatment group:** Boosting type | ['gbdt', 'dart'] |
| | **Outcome model in treatment group:** Maximum tree depth | [3, unlimited] |
| *X-Lasso* | **Outcome model:** penalty strength | $10^{-2}$ to $10^{2}$, evenly spaced on a logarithmic scale (7 steps) |
| | **Tau model:** penalty strength | $10^{-2}$ to $10^{2}$, evenly spaced on a logarithmic scale (7 steps) |
| | **Propensity model:** penalty strength | $10^{-2}$ to $10^{2}$, evenly spaced on a logarithmic scale (7 steps) |
| *X-GBM* | **Outcome model:** Boosting type | ['gbdt', 'dart'] |
| | **Outcome model:** Maximum tree depth | [3, unlimited] |
| | **Tau model:** Boosting type | ['gbdt', 'dart'] |
| | **Tau model:** Maximum tree depth | [3, unlimited] |
| | **Propensity model:** Boosting type | ['gbdt', 'dart'] |
| | **Propensity model:** Maximum tree depth | [3, unlimited] |

| | | |
|---|---|---|
| *R-Lasso* | **Outcome model:** penalty strength | $10^{-2}$ to $10^{2}$, evenly spaced on a logarithmic scale (7 steps) |
| | **Propensity model:** penalty strength | $10^{-2}$ to $10^{2}$, evenly spaced on a logarithmic scale (7 steps) |
| | **R-learner:** penalty strength | $10^{-2}$ to $10^{2}$, evenly spaced on a logarithmic scale (7 steps) |
| *R-GBM* | **Outcome model:** Boosting type | ['gbdt', 'dart'] |
| | **Outcome model:** Maximum tree depth | [3, unlimited] |
| | **Propensity model:** Boosting type | ['gbdt', 'dart'] |
| | **Propensity model:** Maximum tree depth | [3, unlimited] |
| | **R-learner:** Boosting type | ['gbdt', 'dart'] |
| | **R-learner:** Maximum tree depth | [3, unlimited] |
| *Tian* | penalty strength | $10^{-2}$ to $10^{2}$, evenly spaced on a logarithmic scale (7 steps |
| *Causal Forest* | (min_samples_split) minimum number of samples required to split an internal node | [10, 20, 30] |
| | (min_samples_leaf) minimum number of samples required to be at a leaf node | [10, 20, 30, 40, 50, 60] |
| | Maximum tree depth | [2, 3, 7, unlimited] |

Supplementary Table S3: The Python implementations (using the pymer4 package[1]) for the linear mixed-effects logistic regression models (LMMs) used to estimate the marginal odds ratios (ORs), as well as to perform the interaction test. The term “T” denotes the treatment variable (ie, 0=lower PEEP strategy, 1=higher PEEP strategy), “subgroup” denotes the subgroup variable (for instance, the subgroup as identified by the effect models), “peep_tertile” denotes the PEEP tertile variable (ie, 0=PEEP≤8 $cmH_2O$, 1=PEEP 9-11 $cmH_2O$, 2= PEEP ≥12 $cmH_2O$, “trial” denotes the categorical variable for the randomized trial to which the patient belongs, and “pf_ratio” and “peep” denote the terms for the $PaO_2/FiO_2$ and PEEP as continuous variables, “subgroup_mean” denotes the mean of the subgroup variable in each trial, and “subgroup_centered” denotes the subgroup variable centered about the trial-specific mean of the subgroup variable in each trial.

| Model number | Model to .. | Python Implementation |
|---|---|---|
| *1* | … calculate the marginal odds ratios | formula = "mortality_28 ~ T + (1 \| trial)"<br><br>model = Lmer(formula, data=Y, family='binomial') |
| *2* | .. calculate the conditional odds ratios | formula = "mortality_28 ~ Age + RR + T + (1 \| trial)"<br><br>model = Lmer(formula, data=Y, family='binomial') |
| *3* | .. test the HTE among subgroups | formula = "mortality_28 ~ T + subgroup + T:subgroup + (1 \| trial)"<br><br>model = Lmer(formula, data=Y, family='binomial') |
| *4* | .. test the second-order HTE among subgroups and PEEP tertiles | formula = "mortality_28 ~ T + subgroup + peep_tertile + T:subgroup + T:peep_tertile + T:subgroup:peep_tertile + (1 \| trial)"<br><br>model = Lmer(formula, data=Y, family='binomial') |
| *5* | .. test the HTE among subgroups, disentangling within-study and across-study information (to account for potential aggregation bias[2]) | formula = "mortality ~ T + subgroup + T:subgroup_mean + T:subgroup_centered + (1 \| trial)"<br><br>model = Lmer(formula, data=Y, family='binomial') |
| *6* | .. test the HTE for severity scores | formula = "mortality_28 ~ T + severity_score + T: severity_score + (1 \| trial)"<br><br>model = Lmer(formula, data=Y, family='binomial') |

Supplementary Table S4: Variables selected in the forward selection in each fold of the **outer 'leave-one-trial-out' cross-validation** for the modelling procedure using a **causal forest with forward selection**. 'STOPPED' means that in this round, none of the left over candidate variables improved the discrimination for benefit (ie, the AUC-benefit) compared to the previous round, and the forward selection was stopped.

| ***Left-out trial*** | **Round** | **Selected variable** |
|---|---|---|
| ***ALVEOLI*** | 1 | $C_{RS}$ |
| | 2 | Driving pressure |
| | 3 | $FiO_2$ |
| | 4 | Sex |
| | 5 | STOPPPED |
| ***LOVS*** | 1 | $C_{RS}$ |
| | 2 | Sex |
| | 3 | STOPPED |
| ***EXPRESS*** | 1 | $C_{RS}$ |
| | 2 | Tidal Volume |
| | 3 | STOPPED |

Supplementary Table S5: Variables selected in the forward selection in each round of the 'leave-one-trial-out' cross-validation for training the **final model** (ie, using the complete train cohort to train) in the **lower PEEP tertile (using the S-GBM)** and the **higher PEEP tertile (using the causal forest)**. 'STOPPED' means that in this round, none of the left over candidate variables improved the discrimination for benefit (ie, the AUC-benefit) compared to the previous round, and the forward selection was stopped.

(a)

| ***Left-out cohort*** | **Round** | **Selected variable** |
|---|---|---|
| ***Test cohort*** | 1 | Driving pressure |
| | 2 | Pulmonary ARDS (0=no, 1=yes) |
| | 3 | STOPPED |

(b)

| ***Left-out cohort*** | **Round** | **Selected variable** |
|---|---|---|
| ***Test cohort*** | 1 | Driving Pressure |
| | 2 | Tidal Volume |
| | 3 | STOPPED |

Supplementary Figure S1: Schematic overview of the method selection procedure, including the 'outer' and 'nested' leave-one-trial-out cross-validation (LOTO-CV).
*Each candidate method is implemented with, and without forward selection, which is visualized in detail in Figure S2.
**The training of the final models consists of a potential forward selection and hyperparameter optimization, as visualized in Figure S3.

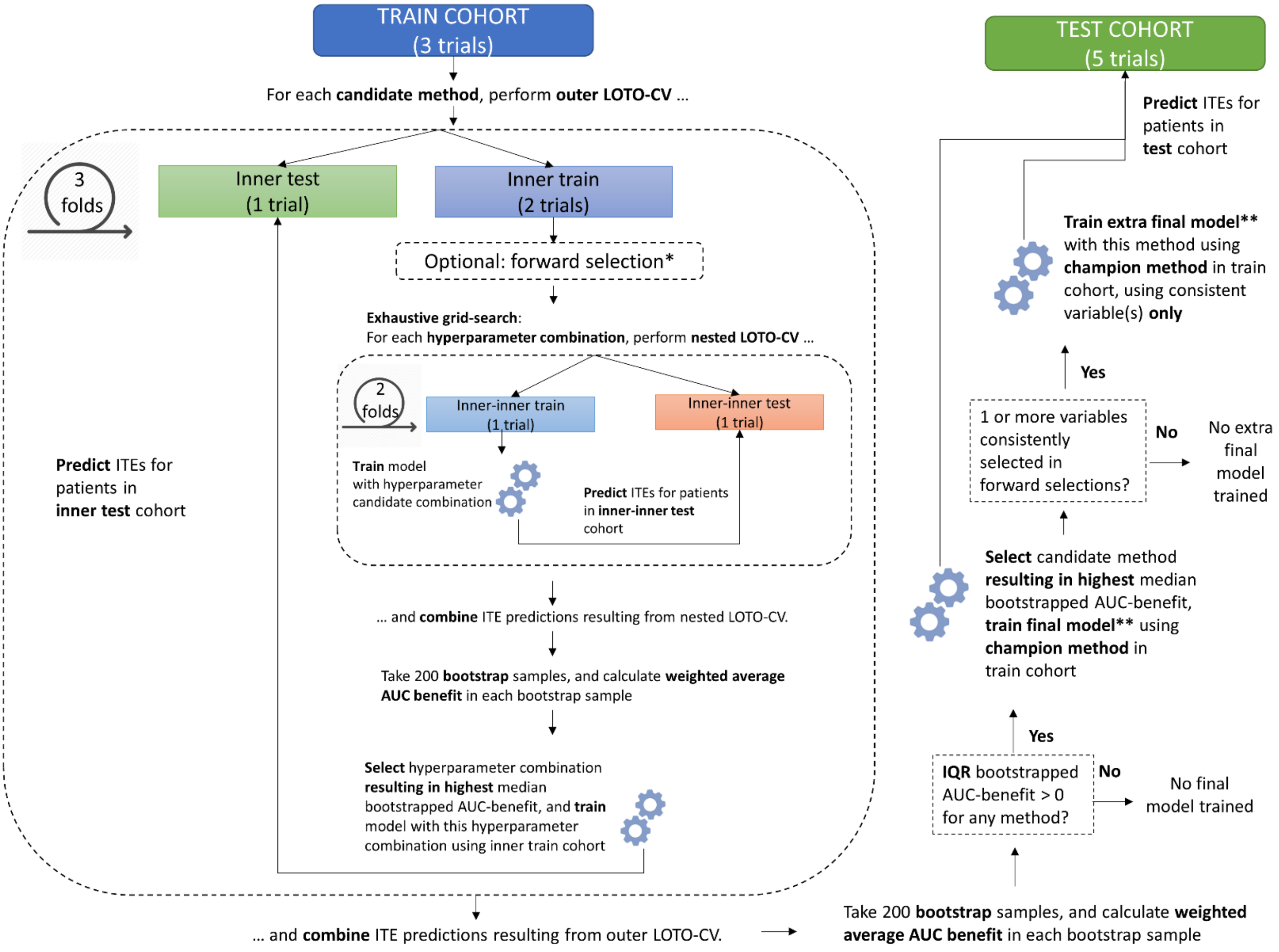

Supplementary Figure S2: Schematic overview of the forward selection procedure.
* If the forward selection procedure is performed for training a final model, the procedure starts with the full train cohort (including three trials), and the LOTO-CVs will consist of three folds, each splitting the (full) train cohort into an inner train cohort (2 trials) and an inner test cohort (one trial).

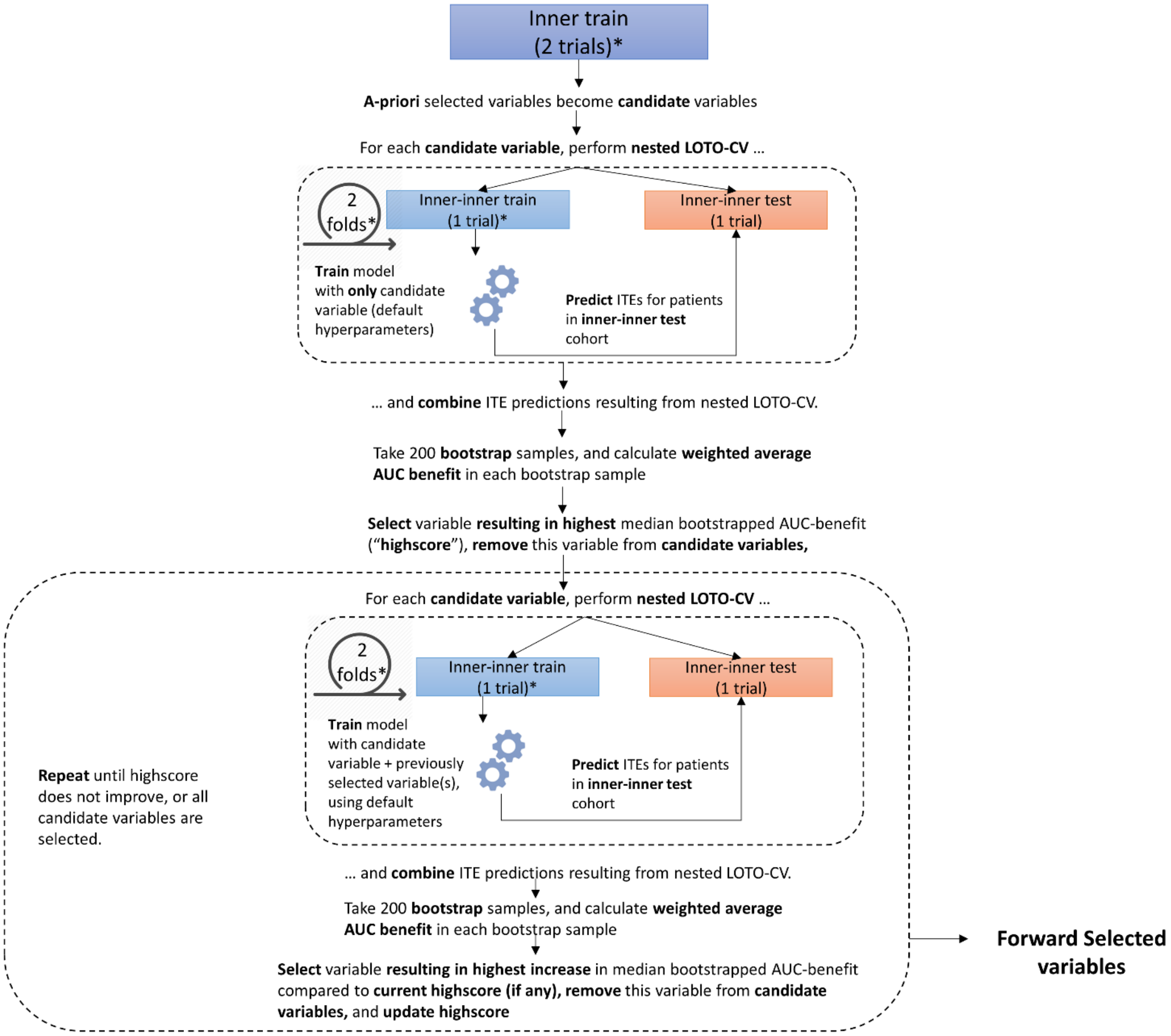

Supplementary Figure S3: Schematic overview of the training of final models.
*if a final model is trained using a method implemented with forward selection, the forward selection is performed using all three trials from the train cohort (see Figure S2). For training the extra final model(s), the consistent variables were selected rather than performing a forward selection procedure.

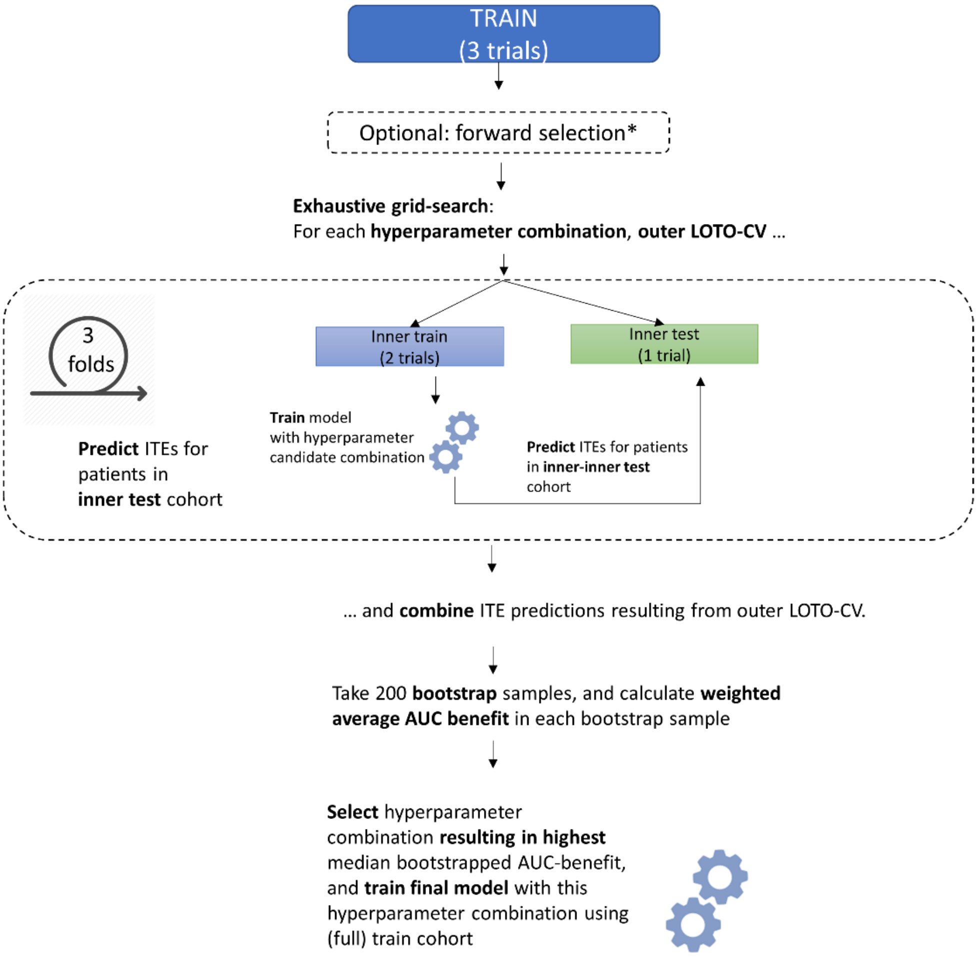

Supplementary Figure S4: Daily ventilatory settings during study days 1-7 in the experimental groups compared to the control groups of the eight included trials.

(a) Mean PEEP levels

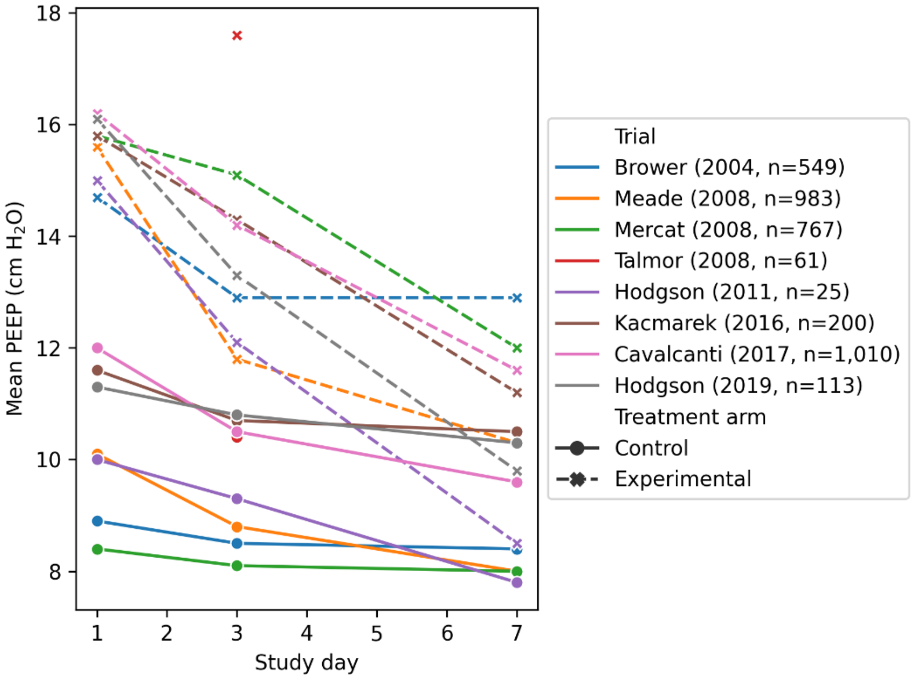


(b) Mean $FiO_2$ levels (this information was not available for the Cavalcanti et al.[3] and Hodgson et al. 2019[4] trials).

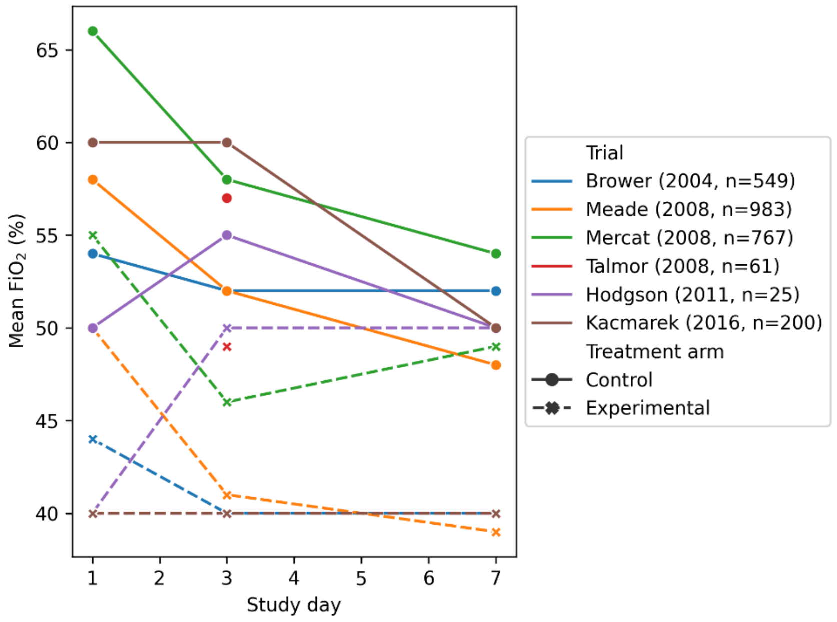

(c) Mean $PaO_2/FiO_2$ levels (this information was not available for the Hodgson et al. 2011[5] trial).

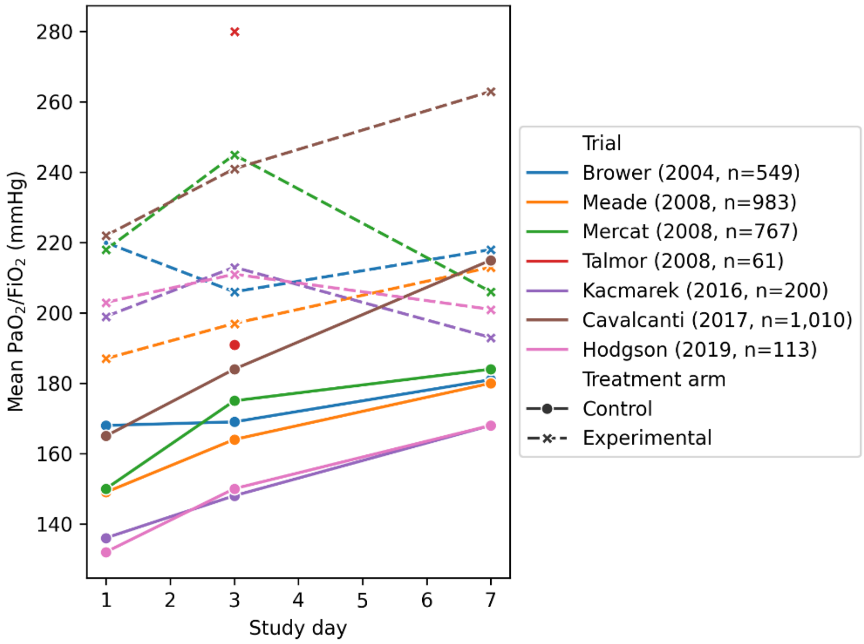

Supplementary Figure S5: Heterogeneity of treatment effect results of 'apparent' validation (ie, models both trained and evaluated in train cohort). Treatment effects of higher vs lower PEEP on the relative, odds ratio scale and the absolute, mortality risk difference scale, plotted for patients from the (**a) ALVEOLI[6] (n=243), (b) LOVS[7] (n=194) and (c) EXPRESS[8] (n=462) trial with baseline PEEP ≤ 8 cm$H_2$O**, classified into the predicted harm, and predicted benefit from high PEEP by **final model 1**. NNT=number needed to treat.

(a) ALVEOLI

| | Lower PEEP group, 28-d mort, n (%) | Higher PEEP group, 28-d mort, n (%) | OR (95% CI) | Mortality reduction, % (95% CI) | NNT | P value for interaction |
|---|---|---|---|---|---|---|
| All patients (n=243) | 18/125 (14.4) | 26/118 (22.0) | 1.68 (0.87; 3.26) | -7.6% (-16.1 to 0.4) | -13 | |
| **Model-based subgroups** | | | | | | <0.001 |
| Predicted harm from high PEEP (ITE≤0, n=114) | 1/66 (1.5) | 19/48 (39.6) | 42.59 (5.44; 333.45) | -38.1% (-50.0 to -25.6) | -2 | |
| Predicted benefit from high PEEP (ITE>0, n=129) | 17/59 (28.8) | 7/70 (10.0) | 0.27 (0.1; 0.72) | 18.8% (7.4 to 30.1) | 5 | |

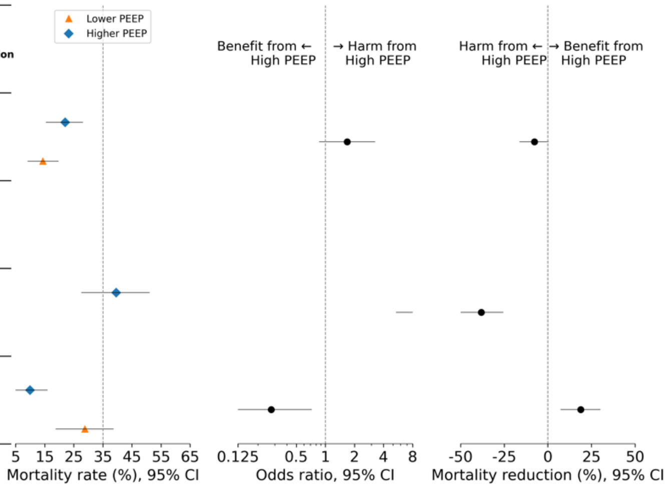


(b) LOVS

| | Lower PEEP group, 28-d mort, n (%) | Higher PEEP group, 28-d mort, n (%) | OR (95% CI) | Mortality reduction, % (95% CI) | NNT | P value for interaction |
|---|---|---|---|---|---|---|
| All patients (n=194) | 21/108 (19.4) | 24/86 (27.9) | 1.6 (0.82; 3.13) | -8.5% (-18.5 to 1.2) | -11 | |
| **Model-based subgroups** | | | | | | <0.001 |
| Predicted harm from high PEEP (ITE≤0, n=111) | 9/70 (12.9) | 17/41 (41.5) | 4.8 (1.88; 12.24) | -28.6% (-42.9 to -14.5) | -3 | |
| Predicted benefit from high PEEP (ITE>0, n=83) | 12/38 (31.6) | 7/45 (15.6) | 0.4 (0.14; 1.15) | 16.0% (-0.7 to 30.4) | 6 | |

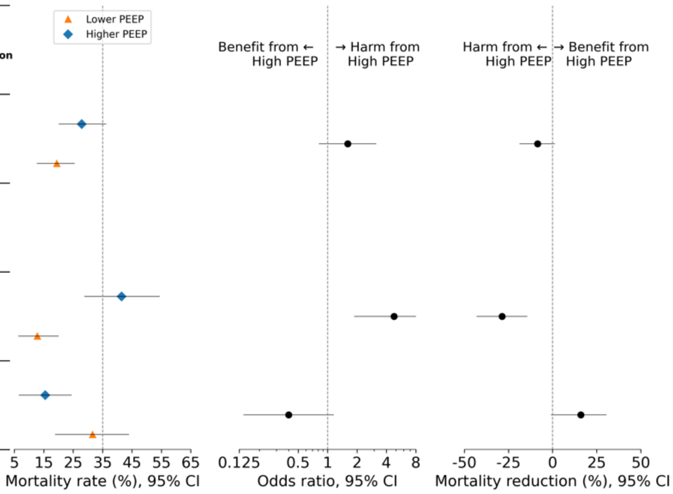

(c) EXPRESS

| | Lower PEEP group, 28-d mort, n (%) | Higher PEEP group, 28-d mort, n (%) | OR (95% CI) | Mortality reduction, % (95% CI) | NNT | P value for interaction |
|---|---|---|---|---|---|---|
| All patients (n=462) | 78/241 (32.4) | 55/221 (24.9) | 0.69 (0.46; 1.04) | 7.5% (1.1 to 14.2) | 13 | |
| **Model-based subgroups** | | | | | | <0.001 |
| Predicted harm from high PEEP (ITE≤0, n=228) | 17/126 (13.5) | 46/102 (45.1) | 5.27 (2.77; 10.02) | -31.6% (-40.8 to -22.0) | -3 | |
| Predicted benefit from high PEEP (ITE>0, n=234) | 61/115 (53.0) | 9/119 (7.6) | 0.07 (0.03; 0.16) | 45.5% (36.6 to 53.3) | 2 | |

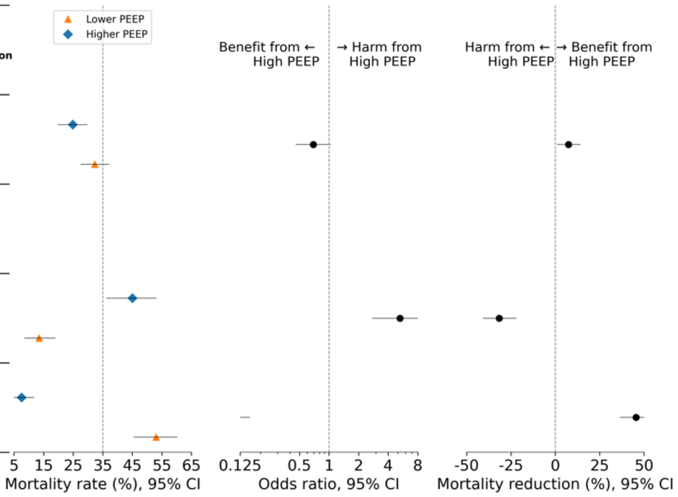


Supplementary Figure S6: Heterogeneity of treatment effect results of ‘apparent’ validation (ie, models both trained and evaluated in train cohort). Treatment effects of higher vs lower PEEP on the relative, odds ratio scale and the absolute, mortality risk difference scale, plotted for patients from the **(a) ALVEOLI[6] (n=243), (b) LOVS[7] (n=194) and (c) EXPRESS[8] (n=462) trial with baseline PEEP ≤ 8 cm$H_2O$**, classified into the predicted harm, and predicted benefit from high PEEP by **final model 2**. NNT=number needed to treat.

(a) ALVEOLI

| | Lower PEEP group, 28-d mort, n (%) | Higher PEEP group, 28-d mort, n (%) | OR (95% CI) | Mortality reduction, % (95% CI) | NNT | P value for interaction |
|---|---|---|---|---|---|---|
| All patients (n=243) | 18/125 (14.4) | 26/118 (22.0) | 1.68 (0.87; 3.26) | -7.6% (-16.1 to 0.4) | -13 | |
| **Model-based subgroups** | | | | | | 0.006 |
| Predicted harm from high PEEP (ITE≤0, n=134) | 6/66 (9.1) | 19/68 (27.9) | 3.88 (1.44; 10.46) | -18.9% (-29.7 to -7.3) | -5 | |
| Predicted benefit from high PEEP (ITE>0, n=109) | 12/59 (20.3) | 7/50 (14.0) | 0.64 (0.23; 1.77) | 6.3% (-4.9 to 18.5) | 15 | |

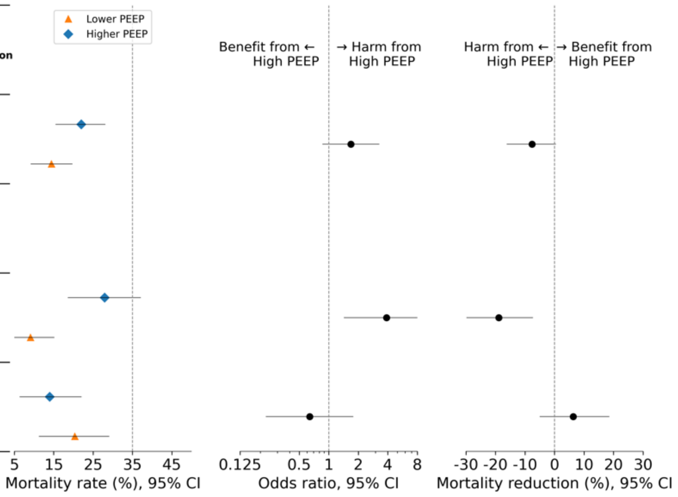

#### (b) LOVS

| | Lower PEEP group, 28-d mort, n (%) | Higher PEEP group, 28-d mort, n (%) | OR (95% CI) | Mortality reduction, % (95% CI) | NNT | P value for interaction |
|---|---|---|---|---|---|---|
| All patients (n=194) | 21/108 (19.4) | 24/86 (27.9) | 1.6 (0.82; 3.13) | -8.5% (-18.5 to 1.2) | -11 | |
| **Model-based subgroups** | | | | | | <0.001 |
| Predicted harm from high PEEP (ITE≤0, n=107) | 6/56 (10.7) | 21/51 (41.2) | 5.83 (2.12; 16.08) | -30.5% (-42.9 to -18.2) | -3 | |
| Predicted benefit from high PEEP (ITE>0, n=87) | 15/52 (28.8) | 3/35 (8.6) | 0.23 (0.06; 0.87) | 20.3% (5.7 to 31.6) | 4 | |

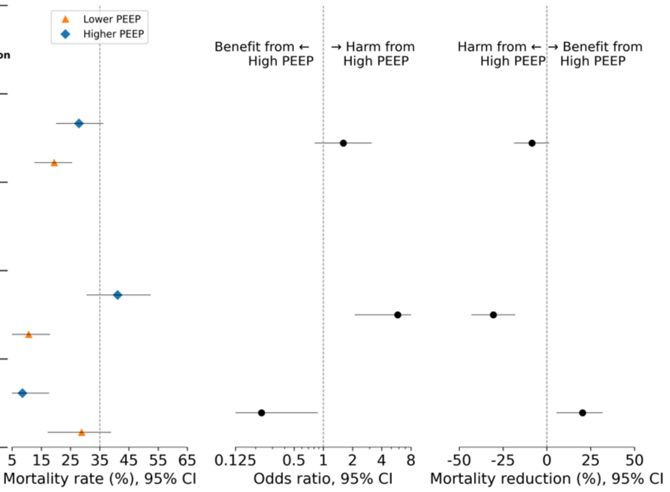


#### c) EXPRESS

| | Lower PEEP group, 28-d mort, n (%) | Higher PEEP group, 28-d mort, n (%) | OR (95% CI) | Mortality reduction, % (95% CI) | NNT | P value for interaction |
|---|---|---|---|---|---|---|
| All patients (n=462) | 78/241 (32.4) | 55/221 (24.9) | 0.69 (0.46; 1.04) | 7.5% (1.1 to 14.2) | 13 | |
| **Model-based subgroups** | | | | | | 0.009 |
| Predicted harm from high PEEP (ITE≤0, n=220) | 33/127 (26.0) | 31/93 (33.3) | 1.42 (0.79; 2.56) | -7.3% (-17.1 to 2.7) | -13 | |
| Predicted benefit from high PEEP (ITE>0, n=242) | 45/114 (39.5) | 24/128 (18.8) | 0.35 (0.2; 0.63) | 20.7% (11.3 to 29.9) | 4 | |

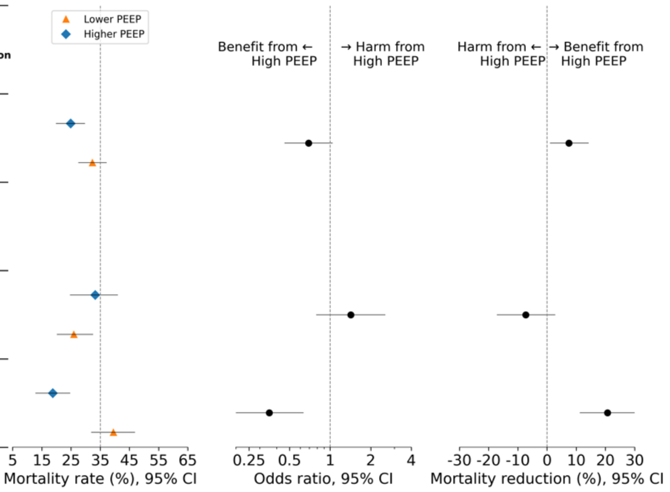

Supplementary Figure S7: Heterogeneity of treatment effect results of ‘apparent’ validation (ie, models both trained and evaluated in train cohort). Treatment effects of higher vs lower PEEP on the relative, odds ratio scale and the absolute, mortality risk difference scale, plotted for patients from the (**a) ALVEOLI[6] (n=141), (b) LOVS[7] (n=481) and (c) EXPRESS[8] (n=136) trial with baseline PEEP ≥ 12 cmH$_2$O**, classified into the predicted harm, and predicted benefit from high PEEP by **final model 3**. NNT=number needed to treat.

(a) ALVEOLI

| | **Lower PEEP group, 28-d mort, n (%)** | **Higher PEEP group, 28-d mort, n (%)** | **OR (95% CI)** | **Mortality reduction, % (95% CI)** | **NNT** | **P value for interaction** |
|---|---|---|---|---|---|---|
| All patients (n=141) | 14/61 (23.0) | 22/80 (27.5) | 1.27 (0.59; 2.76) | -4.5% (-14.8 to 7.1) | -21 | |
| **Model-based subgroups** | | | | | | 0.064 |
| Predicted harm from high PEEP (ITE≤0, $C_{RS}$ ≤ 26.46 cmH$_2$O, n=50) | 3/18 (16.7) | 13/32 (40.6) | 3.42 (0.82; 14.24) | -24.0% (-45.2 to -4.1) | -4 | |
| Predicted benefit from high PEEP (ITE>0, $C_{RS}$ > 26.46 cmH$_2$O, n=91) | 11/43 (25.6) | 9/48 (18.8) | 0.67 (0.25; 1.82) | 6.8% (-5.6 to 21.4) | 14 | |

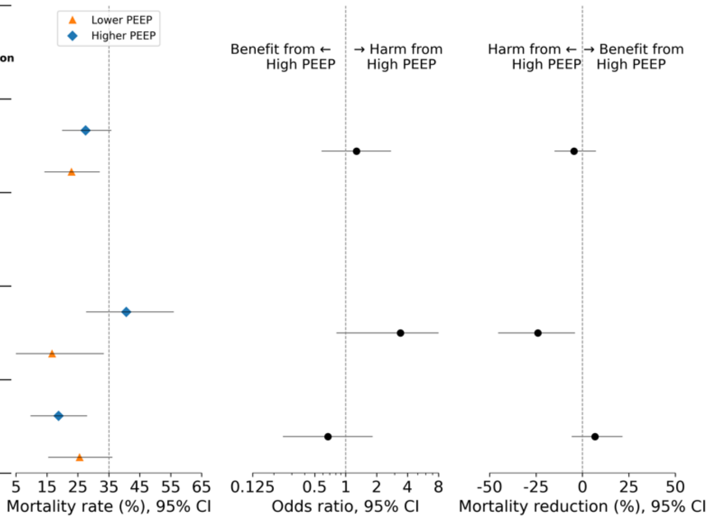


(b) LOVS

| | **Lower PEEP group, 28-d mort, n (%)** | **Higher PEEP group, 28-d mort, n (%)** | **OR (95% CI)** | **Mortality reduction, % (95% CI)** | **NNT** | **P value for interaction** |
|---|---|---|---|---|---|---|
| All patients (n=481) | 91/238 (38.2) | 72/243 (29.6) | 0.68 (0.47; 0.99) | 8.6% (1.4 to 15.6) | 11 | |
| **Model-based subgroups** | | | | | | 0.039 |
| Predicted harm from high PEEP (ITE≤0, n=115) | 20/50 (40.0) | 33/65 (50.8) | 1.55 (0.73; 3.26) | -10.8% (-25.9 to 6.2) | -9 | |
| Predicted benefit from high PEEP (ITE>0, n=366) | 71/188 (37.8) | 39/178 (21.9) | 0.46 (0.29; 0.73) | 15.9% (7.9 to 23.2) | 6 | |

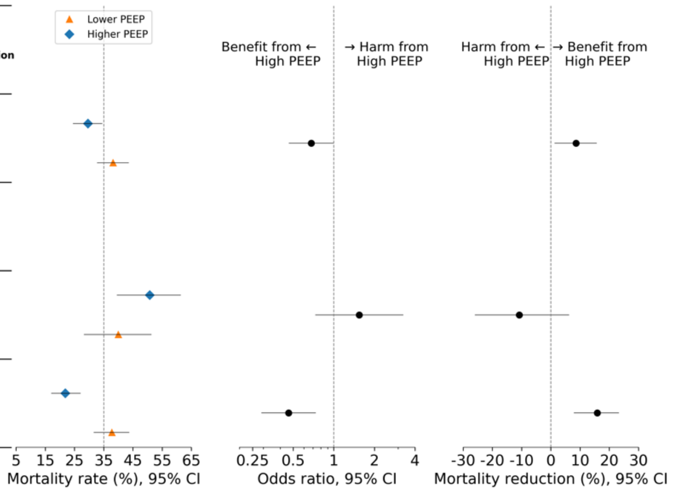

(c) EXPRESS

| | Lower PEEP group, 28-d mort, n (%) | Higher PEEP group, 28-d mort, n (%) | OR (95% CI) | Mortality reduction, % (95% CI) | NNT | P value for interaction |
|---|---|---|---|---|---|---|
| All patients (n=136) | 23/57 (40.4) | 26/79 (32.9) | 0.73 (0.36; 1.47) | 7.4% (-7.5 to 21.0) | 13 | |
| **Model-based subgroups** | | | | | | 0.056 |
| Predicted harm from high PEEP (ITE≤0, n=10) | 1/5 (20.0) | 4/5 (80.0) | 16.0 (0.72; 354.8) | -60.0% (-100.0 to 0.0) | -1 | |
| Predicted benefit from high PEEP (ITE>0, n=126) | 22/52 (42.3) | 22/74 (29.7) | 0.58 (0.27; 1.21) | 12.6% (-2.9 to 27.0) | 7 | |

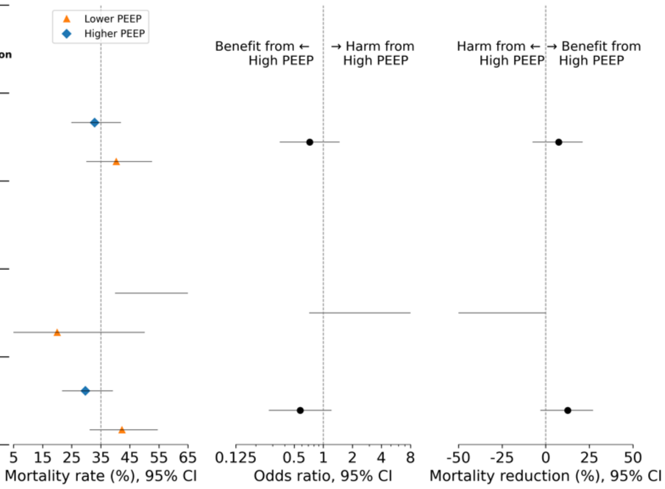


Supplementary Figure S8: Heterogeneity of treatment effect results of 'apparent' validation (ie, models both trained and evaluated in train cohort). Treatment effects of higher vs lower PEEP on the relative, odds ratio scale and the absolute, mortality risk difference scale, plotted for patients from the **(a) ALVEOLI[6] (n=141), (b) LOVS[7] (n=481) and (c) EXPRESS[8] (n=136) trial with baseline PEEP ≥ 12 cmH$_2$O**, classified into the predicted harm, and predicted benefit from high PEEP by **final model 4**. NNT=number needed to treat.

(a) ALVEOLI

| | Lower PEEP group, 28-d mort, n (%) | Higher PEEP group, 28-d mort, n (%) | OR (95% CI) | Mortality reduction, % (95% CI) | NNT | P value for interaction |
|---|---|---|---|---|---|---|
| All patients (n=141) | 14/61 (23.0) | 22/80 (27.5) | 1.27 (0.59; 2.76) | -4.5% (-14.8 to 7.1) | -21 | |
| **Model-based subgroups** | | | | | | 0.064 |
| Predicted harm from high PEEP (ITE≤0, $C_{RS}$ < 26.5 mL/cmH$_2$O, n=50) | 3/18 (16.7) | 13/32 (40.6) | 3.42 (0.82; 14.24) | -24.0% (-45.2 to -4.1) | -4 | |
| Predicted benefit from high PEEP (ITE>0, $C_{RS}$ ≥ 26.5 mL/cmH$_2$O, n=91) | 11/43 (25.6) | 9/48 (18.8) | 0.67 (0.25; 1.82) | 6.8% (-5.6 to 21.4) | 14 | |

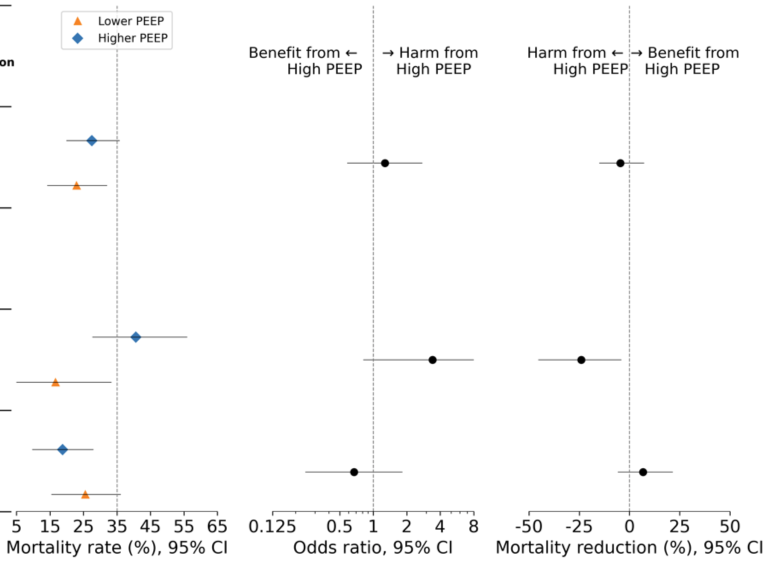

#### (b) LOVS

| | Lower PEEP group, 28-d mort, n (%) | Higher PEEP group, 28-d mort, n (%) | OR (95% CI) | Mortality reduction, % (95% CI) | NNT | P value for interaction |
|---|---|---|---|---|---|---|
| All patients (n=481) | 91/238 (38.2) | 72/243 (29.6) | 0.68 (0.47; 0.99) | 8.6% (1.4 to 15.6) | 11 | |
| **Model-based subgroups** | | | | | | 0.243 |
| Predicted harm from high PEEP (ITE≤0, $C_{RS}$ < 26.5 mL/cmH$_2$O, n=165) | 31/79 (39.2) | 35/86 (40.7) | 1.06 (0.57; 1.98) | -1.5% (-14.7 to 11.0) | -68 | |
| Predicted benefit from high PEEP (ITE>0, $C_{RS}$ ≥ 26.5 mL/cmH$_2$O, n=316) | 60/159 (37.7) | 37/157 (23.6) | 0.51 (0.31; 0.83) | 14.2% (5.2 to 22.2) | 7 | |

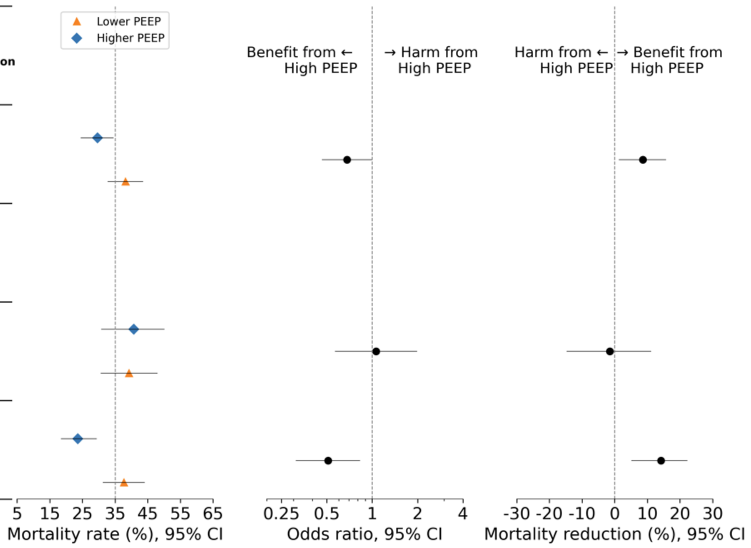


#### (c) EXPRESS

| | Lower PEEP group, 28-d mort, n (%) | Higher PEEP group, 28-d mort, n (%) | OR (95% CI) | Mortality reduction, % (95% CI) | NNT | P value for interaction |
|---|---|---|---|---|---|---|
| All patients (n=136) | 23/57 (40.4) | 26/79 (32.9) | 0.73 (0.36; 1.47) | 7.4% (-7.5 to 21.0) | 13 | |
| **Model-based subgroups** | | | | | | 0.284 |
| Predicted harm from high PEEP (ITE≤0, $C_{RS}$ < 26.5 mL/cmH$_2$O, n=29) | 4/12 (33.3) | 8/17 (47.1) | 1.78 (0.38; 8.23) | -13.7% (-41.4 to 16.7) | -7 | |
| Predicted benefit from high PEEP (ITE>0, $C_{RS}$ ≥ 26.5 mL/cmH$_2$O, n=107) | 19/45 (42.2) | 18/62 (29.0) | 0.56 (0.25; 1.25) | 13.2% (-3.7 to 29.4) | 7 | |

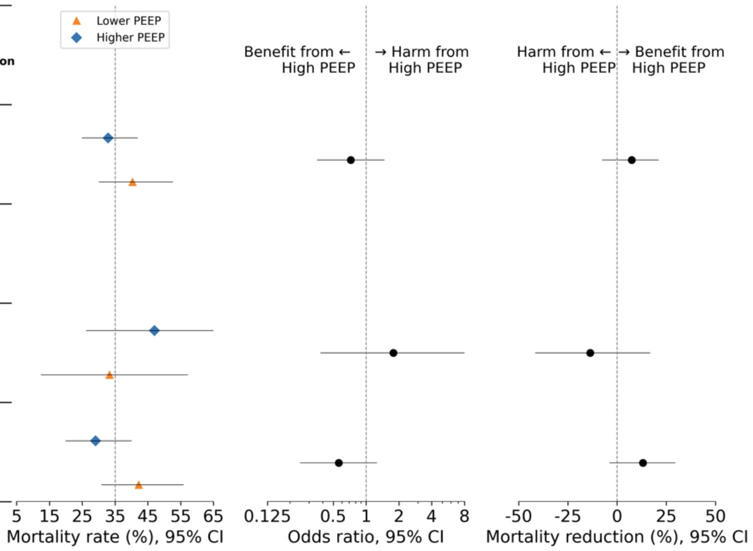

Supplementary Figure S9: Calibration for benefit results, including AUC-benefit, of ‘apparent’ validation (ie, models both trained and evaluated in train cohort). For patients in the **(a) ALVEOLI[6] (n=243), (b) LOVS[7] (n=194) and (c) EXPRESS[8] (n=462) trial with baseline PEEP ≤ 8 $cmH_2O$**, patients are split into four subgroups based on ascending quartiles of the predicted individualised treatment effects (ITEs) predicted by **final model 1**. The predicted ITE distributions are visualised using violin plots next to the observed mortality reductions in each quartile. Error bars indicate 95% CIs.

(a) ALVEOLI (b) LOVS (c) EXPRESS

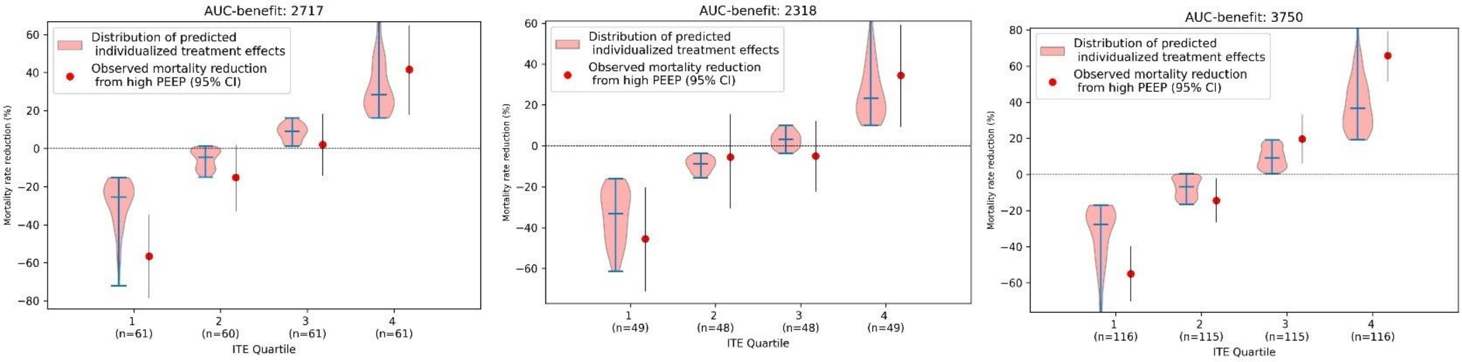


Supplementary Figure S10: Calibration for benefit results, including AUC-benefit, of ‘apparent’ validation (ie, models both trained and evaluated in train cohort). For patients in the **(a) ALVEOLI[6] (n=243), (b) LOVS[7] (n=194) and (c) EXPRESS[8] (n=462) trial with baseline PEEP ≤ 8 $cmH_2O$**, patients are split into four subgroups based on ascending quartiles of the predicted individualised treatment effects (ITEs) predicted by **final model 2**. The predicted ITE distributions are visualised using violin plots next to the observed mortality reductions in each quartile. Error bars indicate 95% CIs.

(a) ALVEOLI (b) LOVS (c) EXPRESS

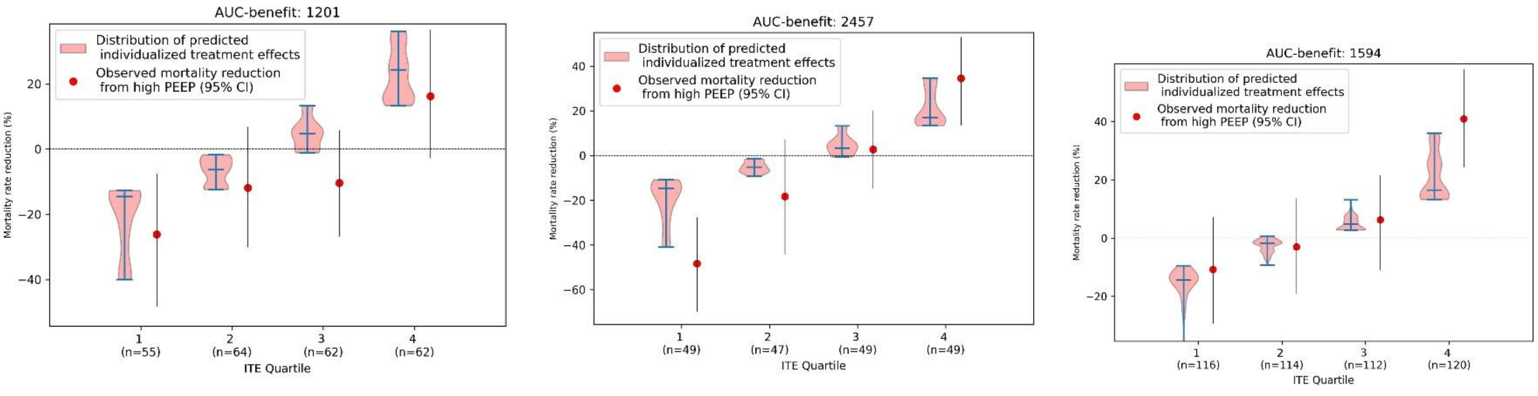

Supplementary Figure S11: Calibration for benefit results, including AUC-benefit, of 'apparent' validation (ie, models both trained and evaluated in train cohort). For patients in the (**a) ALVEOLI[6] (n=141), (b) LOVS[7] (n=481) and (c) EXPRESS[8] (n=136) trial with baseline PEEP ≥ 12 cmH$_2$O**, patients are split into four subgroups based on ascending quartiles of the predicted individualised treatment effects (ITEs) predicted by **final model 3**. The predicted ITE distributions are visualised using violin plots next to the observed mortality reductions in each quartile. Error bars indicate 95% CIs.

(a) ALVEOLI (b) LOVS (c) EXPRESS

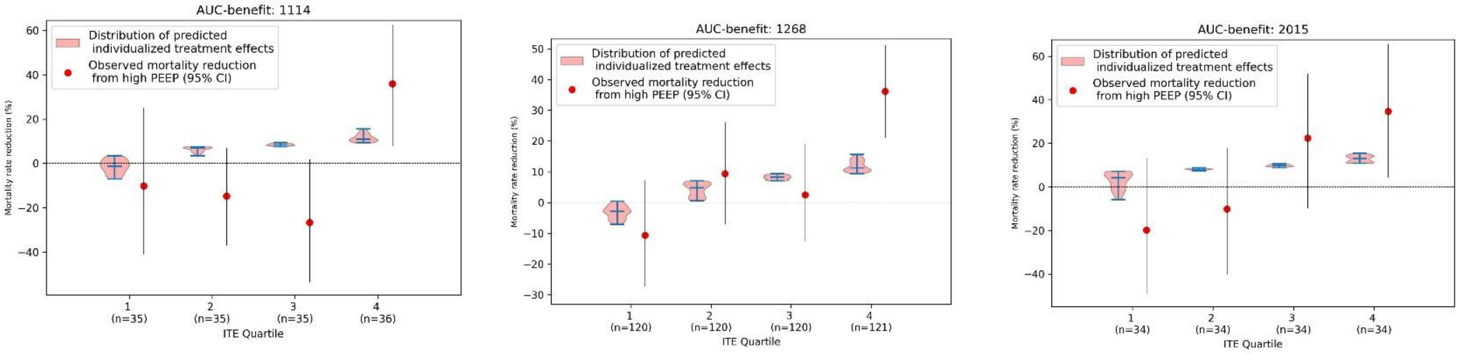


Supplementary Figure S12: Calibration for benefit results, including AUC-benefit, of 'apparent' validation (ie, models both trained and evaluated in train cohort). For patients in the (**a) ALVEOLI[6] (n=141), (b) LOVS[7] (n=481) and (c) EXPRESS[8] (n=136) trial with baseline PEEP ≥ 12 cmH$_2$O**, patients are split into four subgroups based on ascending quartiles of the predicted individualised treatment effects (ITEs) predicted by **final model 4**. The predicted ITE distributions are visualised using violin plots next to the observed mortality reductions in each quartile. Error bars indicate 95% CIs.

(a) ALVEOLI (b) LOVS (c) EXPRESS

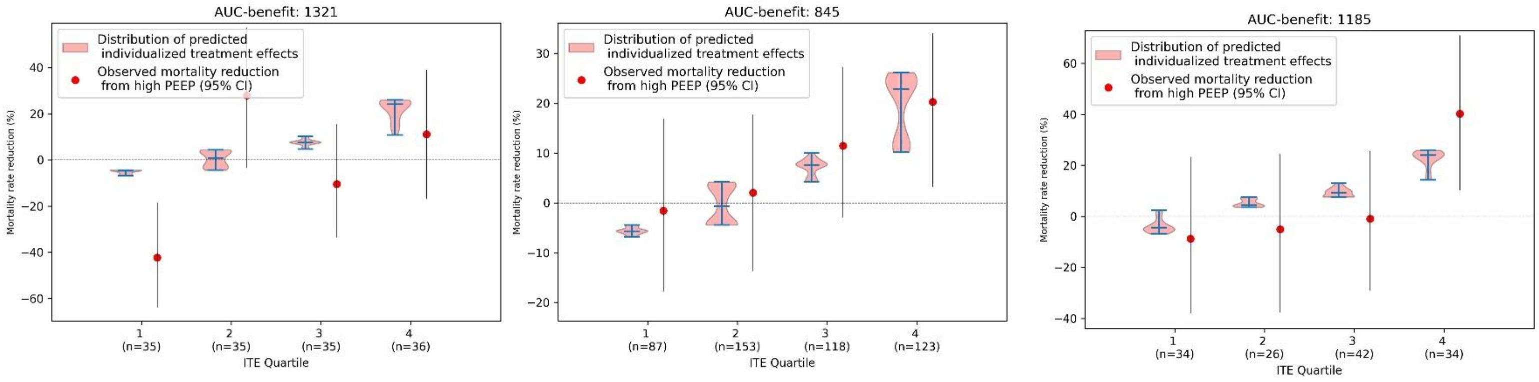

**Appendix part 2:** Definition of the Area under the 'benefit curve' (ie, 'AUC-benefit')

Derivation of AUC-benefit

In this study, we implemented a version of the 'AUC-benefit' which is an update to the definition we proposed in an earlier study, where we now repeatedly split the patients into two groups, not based on a specific individualized treatment effect (ITE) threshold, but based on the patient's rank, after sorting the patients based on ITE.

The calculation of the AUC-benefit comprises the following steps:

1. First, all the patients are ranked based on the predicted ITE, from the lowest to the highest prediction.
2. We divide the patients into two subgroups, with the 25% of the patients with the lowest predicted individualized treatment effects (ie, the $25^{th}$ percentile) in subgroup 1, and the remaining patients in the other subgroup 2.
3. The absolute benefit in terms of 28-day mortality rate reduction (%) is calculated in both subgroups, and the Δ-benefit is defined as the absolute benefit in subgroup 2 minus the absolute benefit in subgroup 1.
4. Then, steps 2 and 3 are repeated 9 times, but then with subgroup 1 consisting the $X^{th}$ percentile of the patients, until the $75^{th}$ percentile, using linearly spaced, equal steps.
5. The resulting Δ-benefit's calculated in steps 2-4 are plotted against the corresponding percentiles, creating the Δ-benefit curve.
6. Finally, the area under the Δ-benefit curve (ie, the 'AUC-benefit') is calculated as the trapezoidal area under this curve. We used Sklearn's 'metrics.auc' function to calculate the AUC-benefit.

AUC-benefit weighted average

As the AUC-benefit metric is for method selection in the ‘outer’ leave-one-trial-out cross-validations (ie, ‘LOTO-CVs’), and also for the forward selection procedure and for the hyperparameter optimization in ‘nested’ LOTO-CVs. Hence, we deal with situations in which individualized treatment effect predictions for different left-out trials, coming from different trained models, need to be jointly evaluated. Simply combining the ITEs predicted during the different cross-validation folds may lead to undesirable effects due to between trial differences in average treatment effects (ie, aggregation bias). Therefore, we for each combination of hyperparameters during the hyperparameter grid search, or candidate feature during the forward selection procedure, we first calculated the AUC-benefits for the predictions resulting from each cross-validation fold, and combined these by calculating the weighted average of these AUC-benefits, using the relative size of the left-out set in each fold as relative to the total size of the data included in the LOTO-CV procedure as the weight.

7 Meade MO, Cook DJ, Guyatt GH, *et al.* Ventilation strategy using low tidal volumes, recruitment maneuvers, and high positive end-expiratory pressure for acute lung

injury and acute respiratory distress syndrome: a randomized controlled trial. *JAMA* 2008; **299**: 637–45.

8 Mercat A, Richard J-CM, Vielle B, *et al.* Positive end-expiratory pressure setting in adults with acute lung injury and acute respiratory distress syndrome: a randomized controlled trial. *JAMA* 2008; **299**: 646–55.
